# TracMyAir: Smartphone-enabled spatiotemporal estimates for inhaled doses of particulate matter and ozone to personalize health outcomes

**DOI:** 10.64898/2026.02.13.26346275

**Authors:** Nicholas F. Lahens, Vlad Isakov, Caroline Chivily, Nadim El Jamal, Antonijo Mrčela, Garret A. FitzGerald, Carsten Skarke

## Abstract

Accurate quantification of individual exposure to air pollutants remains a major challenge in environmental health, as fixed-site monitoring fails to account for mobility, indoor environments, and physiological variability. We deployed TracMyAir, a smartphone-based digital health platform designed to generate time-resolved, personalized exposure and inhaled dose estimates for PM_2.5_ and ozone under real-world conditions. In an exploratory study of 18 adults contributing more than 1,500 participant-hours, the platform integrated smartphone geolocation, regulatory (AirNow) and community-based (PurpleAir) air quality data, building infiltration modeling, microenvironment classification, and wearable-derived physical activity metrics to compute eight tiers of hourly exposure estimates, culminating in individualized inhaled dose. Hourly dose estimates derived from smartphone-and smartwatch-based step counts were concordant (Spearman correlation *p*=0.97–0.98), while heart rate–based estimates yielded greater variability and higher mean values (*p*=0.82–0.92). Exposure explained 51–73% of variance in inhaled dose of PM_2.5_ and 68–84% of ozone, suggesting that physiological-based modeling approaches improve hyperlocal estimates of personal pollutant burden. Substantial inter-and intra-individual variability reflect dynamic microenvironmental transitions and activity patterns. Modeled doses based on regulatory and community sensor networks were strongly correlated (*R*=0.84), with community sensors located closer to participants on average, supporting the feasibility of integrating dense, low-cost monitoring networks. No consistent association was observed between outdoor pollutant levels and neighborhood socioeconomic status in this cohort. These findings demonstrate the feasibility of a scalable, smartphone-centered digital health approach for hyperlocal exposure and inhaled dose modeling. By leveraging ubiquitous consumer devices and existing air quality networks, TracMyAir enables personalized environmental exposure assessment with potential applications in epidemiology, population health, and precision environmental medicine.

## Introduction

Societies become increasingly urban – more than half the world’s population now lives in cities ^1^. Urbanization elevates anthropogenic exposure to air pollutants including particulate matter and ozone from indoor, traffic and industrial emissions. Exposure to ground-level PM_2.5_ and ozone is linked to increased risk of respiratory, cardiovascular, kidney and metabolic ^2^ diseases, premature death, and cancer. While these associations are robust at the population level and supported by mechanistic data from preclinical models and controlled human exposure studies, translating them into individualized risk prediction remains challenging.

Asthma serves as a useful model for examining these complex challenges. Asthma is a chronic lung disorder which causes recurring episodes of bronchospasms commonly triggered by various stimuli including allergens, cold air, exercise and viral respiratory infections ^3^. Development and exacerbation of asthma has been linked to air pollution via oxidative stress, inflammatory responses, airway remodeling, and sensitization to aeroallergens as proposed mechanisms ^4^. This, in part, has been replicated in acute, controlled exposure studies in healthy and asthmatic individuals ^5,6^. Disease susceptibility is modulated by gender ^7^, race and ethnicity ^8^, socioeconomic status ^9,10^, diet ^11^, neighborhood characteristics ^12,13^ and co-morbidities such as obesity, reflux disease or sleep apnea ^14^. A clear association exists between exposure to air pollutants and exacerbations of pre-existing asthma ^15–17^, difficulties with asthma control ^18^ and increased disease risk ^19^. Additionally, asthma exhibits a strong temporal pattern. Up to 75% of asthma patients experience nighttime awakenings due to worsened cough, wheeze and dyspnea ^20–22^, a phenotype associated with poorer disease control, more frequent medication, and higher morbidity and mortality ^23,24^. Circadian clock-dependent diurnal variation in lung function, autonomic tone, hormonal signaling, and airway inflammation contributes to this pattern ^25–30^.

This complexity highlights the challenge of isolating the role of air pollution within the broader network of biological, behavioral, and environmental drivers of asthma. Bridging population-level associations with mechanistic understanding requires temporally resolved, individual-level exposure data capable of aligning pollutant exposure with physiological and pathophysiological processes. Digital health technologies now enable such resolution.

We applied a previously developed efficient and cost-effective mobile phone application called TracMyAir to calculate individual-level air pollution exposures for use in epidemiologic studies and other situations where personal air quality monitoring cannot be performed ^31–34^. The model predicts individual hourly exposures and hourly inhaled doses for PM_2.5_ and ozone while accounting for building-specific infiltration and time spent in different indoor and outdoor microenvironments ^31^.

The objectives for the current study were i) to assess feasibility of TracMyAir in a real-world setting, and ii) to discern the difference between personal exposure to PM_2.5_ and ozone versus personal inhaled dose of PM_2.5_ and ozone.

## Materials and methods

### Study Setup

The air quality assessments of the present study were part of the broader chronobiome study designed to examine time-and age-specific relationships in phenomic and multiomics outputs in a cohort of apparent healthy adults ^35^. We extended this framework by piloting the collection of exposure metrics to PM_2.5_ and ozone under real-world conditions. The study included 18 (non-smoking) adult participants (1563 participant-hours) recruited in the Philadelphia metropolitan area of Pennsylvania (PA). Written informed consent was given by all participants prior to enrollment, and the clinical study was approved by the University of Pennsylvania Institutional Review Board (Federalwide Assurance FWA00004028; IRB Registration: IORG0000029) in compliance with the guidance issued by the International Conference on Harmonization (ICH) harmonized tripartite guidelines: E6 Guideline for Good Clinical Practices. The study protocol was registered as ClinicalTrial NCT04225442. The exposure assessment for each participant was set for a minimum of three consecutive days and up to nine days between March 2021 and August 2021. Study data were collected and managed using REDCap electronic data capture tools hosted at the University of Pennsylvania ^36,37^. REDCap (Research Electronic Data Capture) is a secure, web-based software platform designed to support data capture for research studies, providing 1) an intuitive interface for validated data capture; 2) audit trails for tracking data manipulation and export procedures; 3) automated export procedures for seamless data downloads to common statistical packages; and 4) procedures for data integration and interoperability with external sources.

### Air Quality Monitoring Network

Participants were contacted prior to device deployment to confirm their ability to comply with monitoring. Before deployment, the study team configured the iPhone health app on the study-provided iPhone (iPhone SE 2020, 64 GB on Verizon smartphone data plan) with the participant’s height, weight, first and last initial, date of birth, and sex. This information was then imported into the TracMyAir App. The study iPhone with protective casing (Otterbox Defender), study Apple watch (Series 3 in Space Gray Aluminum Case with Black Sport Band, 42mm case size), external battery (Anker PowerCore 10000 Portable Charger), two study PurpleAir PM_2.5_ air monitors (model PA-II-SD), and a prepaid return shipping label were sent to each participant. The participants individually met virtually with the study team to finish device set up. Participants supplied additional information regarding blood type, Fitzpatrick skin type, wheelchair necessity, and use of medications that affect heart rate. The study team instructed the participants to enter this information into the iPhone Health App and the participants showed the study staff the screen of the iPhone to confirm proper data input. The study team then instructed the participants to complete the “Home Operating Conditions” (e.g. ventilation), “Microenvironments” (e.g. work and home boundaries, in-vehicle time), and “Home Characteristics” (e. g. size and type of home, year built) categories in the TracMyAir App to personalize data collection ^31^. Participants were instructed to keep the iPhone and Apple watch charged, and with them always during the three-day observation period. To activate PM_2.5_ data collection through the PurpleAir monitors, the study team instructed the participants to set up one of the monitors on an elevated surface, such as a nightstand, away from a window. The study team guided the participants through set up using instructions obtained from the PurpleAir website. The second PurpleAir monitor was deployed outdoors, if the participants had access to an outdoor space with an outlet, and access to internet [though the outdoor devices were provided, none of the participants were able to set them up for data collection]. Participants confirmed location of the monitor with the study team via video footage and the study team ensured that the monitor remained in the same location throughout the monitoring period. To register the devices on the manufacturer’s website, a geolocation coordinate point was chosen in a public location near each participant’s address to maintain privacy. The study team conducted virtual meetings with each participant twice daily during the three-day monitoring period to facilitate compliance with the data collection procedure — once in the morning and once in the evening. Within the TracMyAir App, data on “steps from iPhone only”, “steps from Apple watch”, and “heart rate from Apple watch” was collected corresponding to the AirNow regulatory monitoring network and PurpleAir monitoring network. Once the monitoring period was complete, participants returned the devices by mail to the study team. TracMyAir data files were downloaded from the study iPhone through the iTunes computer application.

### Overview of iPhone Application (TracMyAir)

The TracMyAir exposure model is an iOS application for the iPhone smartphone that determines individual-level exposure metrics for PM_2.5_ and ozone ^31–34^. TracMyAir can be operated in two different modes: single-location model and location-logging mode. For the single-location mode, TracMyAir uses the current location of the smartphone to determine daily 24-h average exposures. For the location-logging mode, TracMyAir uses the location history of the smartphone to determine 1-h average exposures. TracMyAir includes a microenvironment classification model based on time-resolved smartphone geolocations, and inhaled ventilation models based on physical activity data from smartphone and smartwatch accelerometers and heart rate sensors. TracMyAir calculates values for several tiers of exposure metrics for use in the epidemiologic analyses. This current study used the location-logging mode.

TracMyAir determines values for eight tiers of individual-level exposure metrics for ambient PM_2.5_ and ozone (Figure 1). The exposure metrics include: measured outdoor concentrations from nearby monitors (Tier 1), three metrics related to PM_2.5_ and ozone infiltration into buildings (Tier 2: residential air exchange rates, Tier 3: infiltration factors, Tier 4: indoor concentrations), three metrics that account for time spent in different indoor and outdoor locations (Tier 5: personal exposure factor, Tier 6: time spent in microenvironments, Tier 7: exposures), and a metric that accounts for physical activity (Tier 8: inhaled dose). The importance of multiple tiers of exposure metrics for epidemiological studies was highlighted in the National Academy of Sciences Report “Health Risks of Indoor Exposure to Particulate Matter” ^38^ and demonstrated in epidemiological studies that used population-level exposure metrics ^39^.

**Figure 1.**
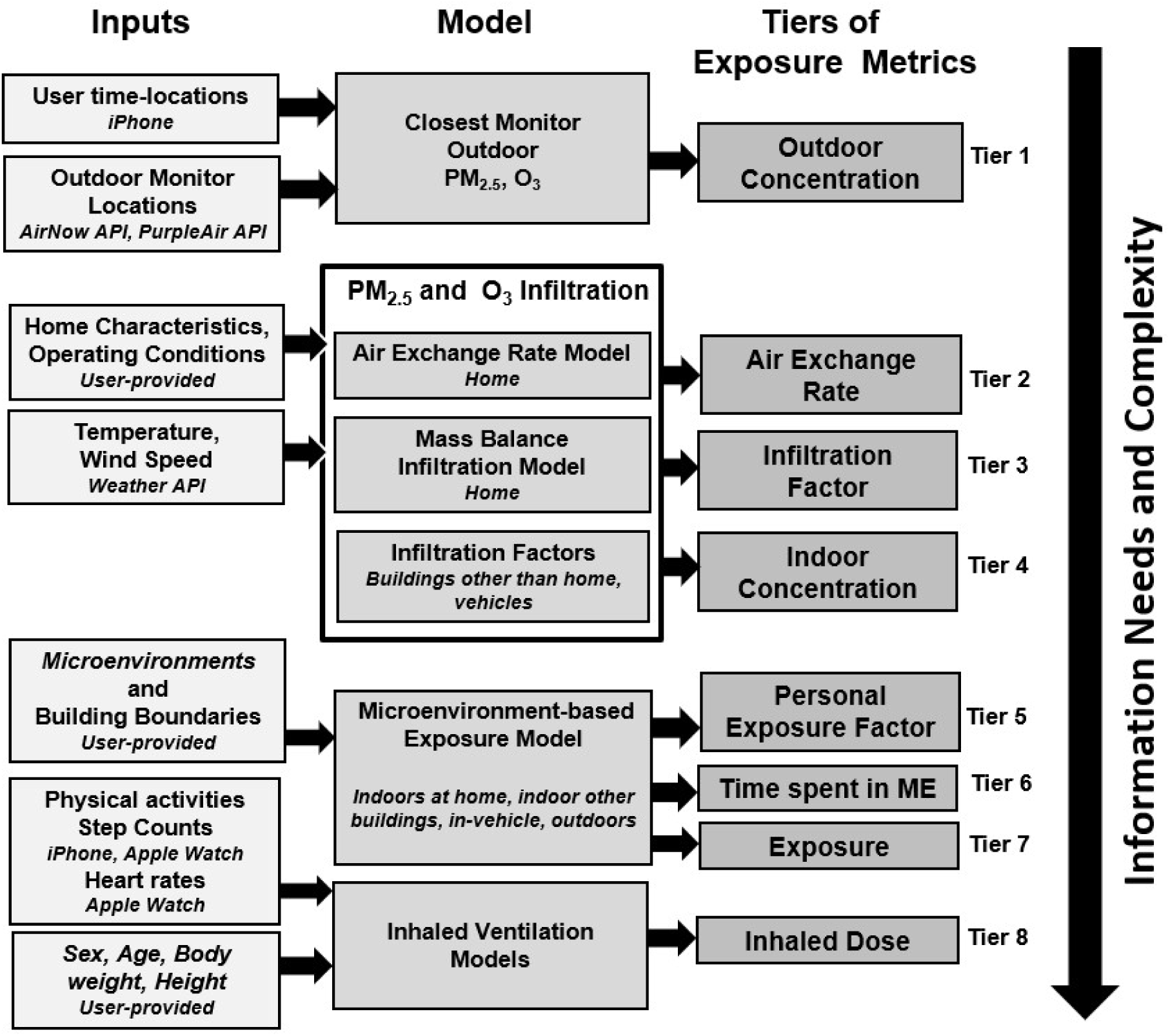
TracMyAir conceptual flow diagram Conceptual flow diagram and tiers to derive exposure metrics predicted by TracMyAir. Tier 1 is a measured exposure metric, whereas Tiers 2-8 are modeled. The application first collects the location history of the smartphone, and then calculates 1-h average exposure metrics based on the location data from the past 24 h.

The TracMyAir input includes outdoor air PM_2.5_ and ozone concentrations, building characteristics and operating conditions, weather, time-locations, and physical activity information. The modeling algorithms include a residential air exchange rate (AER) model, building infiltration model, geolocation-based microenvironment tracker model, microenvironment-based exposure model, and activity-based inhaled ventilation model. The iOS application was written using Swift programming language (version 4.2.1; Apple Inc., Cupertino, CA, USA) and the Xcode Integrated Development Environment. TracMyAir generated 1-h averages for the 8 tiers of exposure metrics for PM_2.5_ and ozone (Figure 1). The tiers have increasing levels of complexity and information needs. Tier 1 is a measured exposure metric, whereas Tiers 2-8 are modeled. The application first collects the location history of the smartphone and then calculates 1-h average exposure metrics based on the location data from the past 24 h.

TracMyAir output variables are provided in **Error! Reference source not found.**, **Error! Reference source not found.**, **Error! Reference source not found.**, **Error! Reference source not found.**, and **Error! Reference source not found.**.

### Data processing and quality control

Study participants exported data from the TracMyAir app twice daily. Each data export produced multiple tabular text files containing measurements collected from the three monitoring station networks (AirNow, PurpleAir, OpenAQ), dosage estimates derived from the three activity measures (Apple watch heart rate, Apple watch step count, iPhone step count), heart rate measurements, and iPhone GPS locations. All files containing PM_2.5_-and ozone-related measurements were merged within each participant. There are cases where multiple input files contained measurements from the same time stamps (due to overlapping data collections), which created duplicates in the merged data table. These are the result of overlaps in the 24-hour measurements periods between successive data exports. To resolve these duplicate time stamps, only those measurements from the most recent data exports were retained for downstream analysis and visualization. This same duplicate resolution method was also applied to the merged heart rate measurements.

Next, to facilitate downstream analyses, hourly measurements were categorized by microenvironment, activity level, and time of day. Briefly, for each measurement the TracMyAir app reports the percentage of the hour the participant spent in each microenvironment. If a participant spent 60% or more of an hour in a given microenvironment the measurement was categorized as coming from the corresponding microenvironment, otherwise the measurement was categorized as coming from a “Mixed” microenvironment. Similarly, TracMyAir reports the percentage of the hour spent at one of four possible activity intensity levels: sedentary, light, moderate, or vigorous. Once again, measurements where participants spent at least 60% of their time at a given intensity level were assigned that intensity, otherwise they were assigned a “Mixed” intensity level. This assignment was repeated for each of the three metrics used by TracMyAir to estimate intensity levels (heart rate and the two step counts). Lastly, each measurement was classified as either day (06:00-22:00) or night (22:00-06:00) based on the wall clock time of the time stamp. All preprocessing steps were applied uniformly across participants.

The 2023 Area Deprivation Index (ADI) rankings mapped to 14-digit FIPS codes from all U.S. states were obtained from the download portal of The Neighborhood Atlas ^40^ website ^41^ (v4.0.1). Note, the 14-digit FIPS (Federal Information Processing Standard) codes uniquely identify ‘census block groups’, geographical units defined by the United States Census Bureau. The tigris R package ^42^ (v2.2.1) was used to retrieve U.S. Census Bureau shapefiles which map census block groups to their geographical locations. Next, the sf R package ^43,44^ (v1.0-21) was used to identify the census block group overlapping the GPS coordinate of each PM_2.5_ and ozone monitoring station. This created a mapping of ADI rank to census block group to monitoring station. This mapping was also used to identify the state/region for each monitoring station

### Statistical software

All data processing, statistical analyses, and figure generation were conducted in R (v4.4.1) using base R packages, tidyverse packages (readr [v2.1.5], dplyr [v1.1.4], tidyr [v1.3.1], purr [v1.0.2], lubridate [v1.9.3], broom [v1.0.6], ggplot2 [3.5.1]), the gt package (v0.11.1), the ggpubr package (v0.6.1), and the patchwork package (1.3.0). Specific statistical tests and summary measures used in this study are specified in the figure legends, tables, and text where their results are reported.

## Results

### Remote sensing domain centered on eastern Pennsylvania and adjacent states

The location-logging version of TracMyAir was used to determine hourly PM_2.5_ and ozone exposures for the n=18 study participants. The remote sensing domain encompassed a large geographic area approximately 200 km x 200 km. The domain included 16 airports which provide hourly meteorological observations needed for exposure calculations. The smaller remote sensing sub-domain centered in Philadelphia included three airports: KLOM (Wings Field Airport), KPNE (Northeast Philadelphia Airport), and KPHL (Philadelphia International Airport) (**Error! Reference source not found.**). There were 5 AirNow monitoring stations for ozone, 7 AirNow monitoring stations for PM_2.5_, and 13 community-embedded outdoor Purple Air PM_2.5_ monitors in this sub-domain. Across the remote sensing domain on average (±SD), 4±3 stations per participant provided data for estimating exposure metrics. The majority of stations (maximum of n=8) was located in eastern Pennsylvania, while a few were in New Jersey (maximum of n=1), Delaware (maximum of n=3), Maryland (maximum of n=3), Virginia (maximum of n=2) and Washington DC (maximum of n=3) (**Error! Reference source not found.*a***). This network of stations was mostly populated by AirNow sensors, followed by community-deployed PurpleAir sensors and OpenAQ. Notably, 2526 and 1903 hourly measurements were contributed, respectively, by AirNow and PurpleAir with a smaller number of 928 hourly measurements provided by OpenAQ (**Error! Reference source not found.*b***). Distributions of measurements collected from each station and participants per station are shown in **Error! Reference source not found.*c*** and ***d***, respectively. OpenAQ was dropped from plots and tables due to incomplete coverage across study participants. Participant-performed data collection revealed that sensing was intermittently interrupted for several participants, as visible, for example, for participant ID#3, #6, #11 and #18, and that sensing periods differed between participant, shorter for ID#5 and longer for ID#10 and #11 (**Error! Reference source not found.**).

Though the study design planned for two PurpleAir sensors to monitor each participant’s indoor and outdoor PM2.5 levels, remote deployment for the indoor sensor was only achieved in n=3 cases and none for the outdoor sensor (frequent reasons included apartment without access to outdoor space, or outdoor space without access to an outlet to power the sensor). This underscores the challenges of adopting IoT (Internet of Things) devices as digital health technologies with participants performing data acquisition.

*Figure 1* illustrates the TracMyAir modelling framework with geocoded input parameters to calculate eight outputs defined as: personal outdoor concentrations (C_out_, Tier 1), residential air exchange rates (AER, Tier 2), infiltration factors (F_inf_home_, Tier 3), indoor concentrations (C_in_home_, Tier 4), personal exposure factors (F_pex_, Tier 5), time spent in ME (T_ME_, Tier 6), exposures (E, Tier 7), and inhaled doses (D, Tier 8). Central site air quality observations (Tier 1, Figure 1) are commonly used as a surrogate for exposure in epidemiologic studies of air pollution. However, depending on the study design, the central site monitor could be far away from the participant study home and work locations. A critical benefit of the TracMyAir mobile app is that by using the nearest PM_2.5_ and ozone monitor data each hour, TracMyAir accounts for a participant’s mobility and for the spatiotemporal variability of outdoor PM_2.5_ and ozone concentrations. To illustrate the importance of time-resolved exposure characterization, we selected a time-course of PM_2.5_ and ozone for a representative study participant. Figure 2 shows distributions of outdoor concentrations and exposures for the different microenvironments of “Indoors”, “Outdoors”, “Vehicle”, and “Mixed”. The figure shows some key characteristics of the time series: i) a rapid hourly change in PM_2.5_ and ozone concentrations when the participant transitions between different microenvironments, ii) an exposure profile which is typically lower indoors, and iii) changes in exposure follow the time-specific pattern of outdoor concentrations. All participant-level exposures over time are shown in **Error! Reference source not found.**. These exposure profiles noticeably show only few stretches of outdoor or mixed instances, an observation likely driven by the COVID-19 pandemic where participants spent less time in these microenvironments. We summarize more detailed findings for the microenvironment-based (Tiers 1-7) modelling of time-specific, geocoded exposure metrics for PM2.5 and ozone in the Supplementary Results.

**Figure 2.**
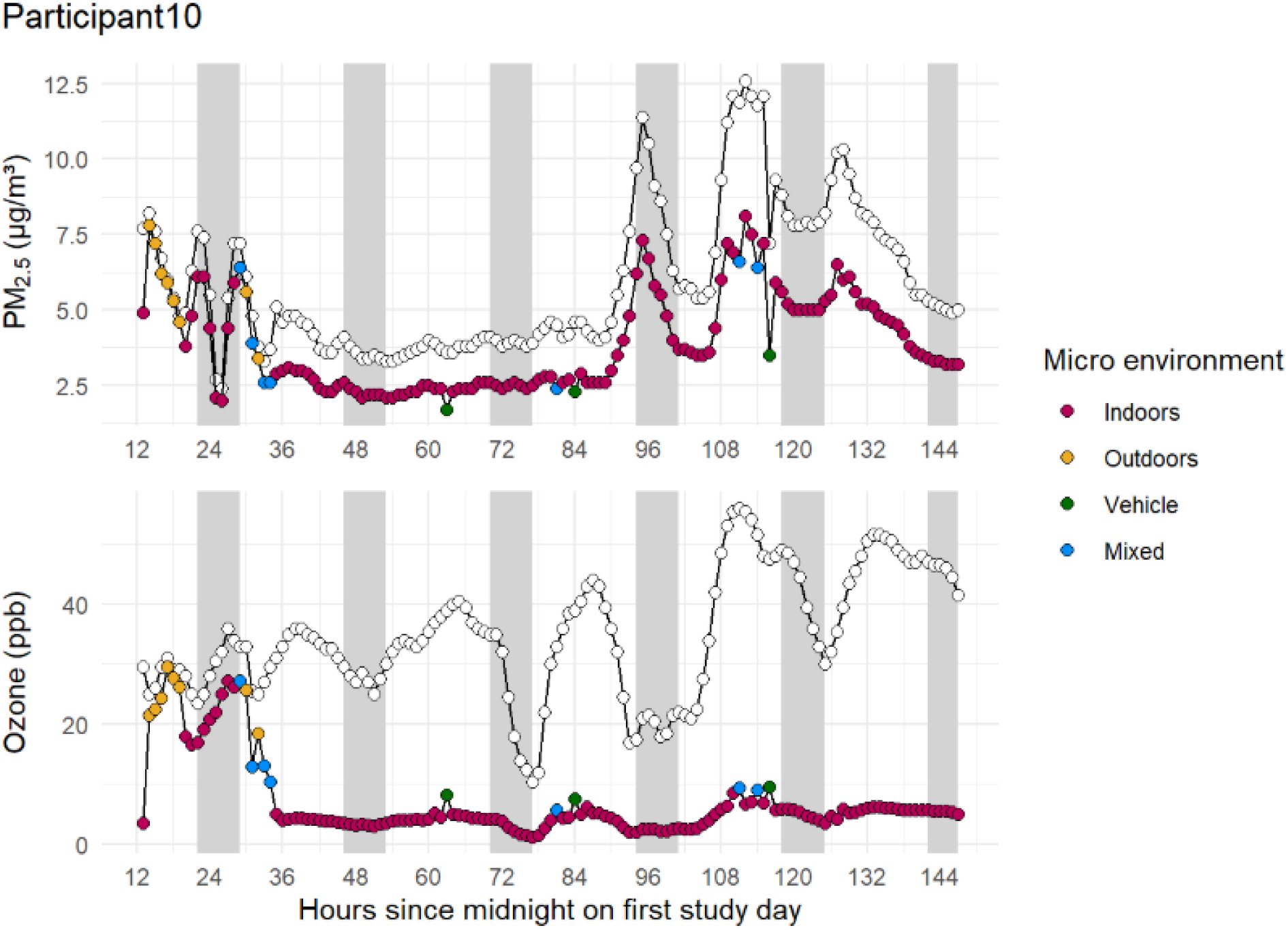
Hyperlocal exposure dynamics of PM_2.5_ and ozone Hyperlocal exposure dynamics of PM_2.5_ and ozone (Tier 7) using AirNow for a single participant navigating through indoor (red circle), outdoor (yellow circle), vehicle (green circle) and mixed (blue circle) microenvironment in comparison to raw ambient outdoor levels of PM2.5 and ozone (white circles). Data panels for the remainder of study participants are shown in **Error! Reference source not found.**

### Modeling time-specific, personalized inhaled dose of PM2.5 and ozone (Tier 8)

To determine hourly inhaled dose as the highest tier of exposure metrics (Tier 8, Figure 1), the TracMyAir model accounted for variations in personal physical activity levels in order to calculate estimates for hourly inhaled doses of PM_2.5_ and ozone. The TracMyAir model has three options for dose calculations which are based on different methods to estimate the minute ventilation parameter using: 1) step counts from the smartphone, 2) step counts from the Apple Watch, and 3) heart rates from the Apple Watch. Hourly inhaled doses of PM_2.5_ and ozone based on AirNow inputs were similar on the cohort level for iPhone-based (1.0±0.8 µg/m^2^ and 2.8±4.0 µg/m^2^, respectively) and Apple Watch-based step counts (1.1±0.8 µg/m^2^ and 3.1±4.4 µg/m^2^, respectively). The Apple Watch-based heart rate, however, provided higher variability and higher mean values of hourly inhaled dose estimates (1.9±1.6 µg/m^2^ and 5.6±7.7 µg/m^2^ for PM_2.5_ and ozone, respectively, **Error! Reference source not found.*a***). The PurpleAir-derived estimates for hourly inhaled doses of PM_2.5_ were comparable. Spearman correlations showed strong concordance for dose estimates based on smartphone-and Apple Watch-derived step counts (*p*=0.97–0.98). Dose estimates based on heart rate compared to the step count-derived were weaker (*p*=0.82–0.92, **Error! Reference source not found.*b***). The higher variability of the heart rate-based hourly inhaled dose is likely attributable to individual differences in the resting heart rate, ranging between 60 and 100 bpm in apparently healthy volunteers ^35,45,46^, and in the intra-individual cardiovascular responses to physical activity. The present version of TracMyAir does not provide the step count as exportable data feature thus not allowing a direct comparison between step count and heart rates.

Distributions of hourly inhaled dose estimates for PM_2.5_ and ozone for each study participant, ranged from 0.4±0.3 µg/m^2^ to 2.2±0.6 µg/m^2^ and from 0.5±0.2 µg/m^2^ to 12.3±9.0 µg/m^2^, respectively, for phone-based step counts. Of similar range were the Apple Watch-based step count-based readouts for hourly inhaled doses (range from 0.4±0.2 µg/m^2^ to 2.3±0.7 µg/m^2^ and from 0.5±0.2 µg/m^2^ to 13.4±9.9 µg/m^2^, respectively). The Apple Watch-based heart rate model produced a broader range from 0.7±0.3 µg/m^2^ to 4.0±1.4 µg/m^2^ and from 0.5±0.3 µg/m^2^ to 17.4±14.2 µg/m^2^, respectively (**Error! Reference source not found.**). These outputs were using AirNow data as input. Estimates were comparable for the PurpleAir-derived hourly inhaled doses of PM_2.5_ (range from 0.4±0.2 µg/m^2^ to 2.0±0.6 µg/m^2^ for phone step counts, 0.2±0.2 µg/m^2^ to 2.1±0.7 µg/m^2^ for Apple Watch step count, and 0.3±0.2 µg/m^2^ to 3.7±1.3 µg/m^2^ for Apple Watch heart rate). Similarly to other tiers of exposure metrics, the hourly inhaled doses vary considerably among participants, a result of temporal variations in personal activity, individual home conditions, and other factors such as time spent in different microenvironments, and meteorological conditions (**Error! Reference source not found.**).

### Variance explained in hourly inhaled doses of PM_2.5_ and ozone

Inhaled doses of PM_2.5_ are inherently related to PM_2.5_ exposures. This relationship, however, is modulated largely by the microenvironment and the level of physical activity. With the AirNow network, 51% of the variance in inhaled hourly doses of PM_2.5_ was explained by exposure to PM_2.5_ when using heart rate as proxy to estimate physical activity. This increased to 73% when step counts were used as proxy (**Error! Reference source not found.**). PurpleAir-based correlations were similar (**Error! Reference source not found.**). For ozone, variance in hourly inhaled dose was explained to a larger degree by exposure, 68% for heart rate as proxy and 84% for step counts (**Error! Reference source not found.**).

### Dynamic directional relationships between PM_2.5_ hourly inhaled dose, heart rate & level of physical activity

Given the interdependency of physical activity, heart rate and inhaled PM_2.5_ dose in our data set and modeling, highly correlated features are expected. Consequently, hourly estimates of inhaled doses of PM_2.5_ tracked with mean heart rate and physical activity shown for single participants with small and large inter-day variability in these relationships (Figure 3). Participant-level plots are shown in exposures over time are shown in **Error! Reference source not found.**.

**Figure 3.**
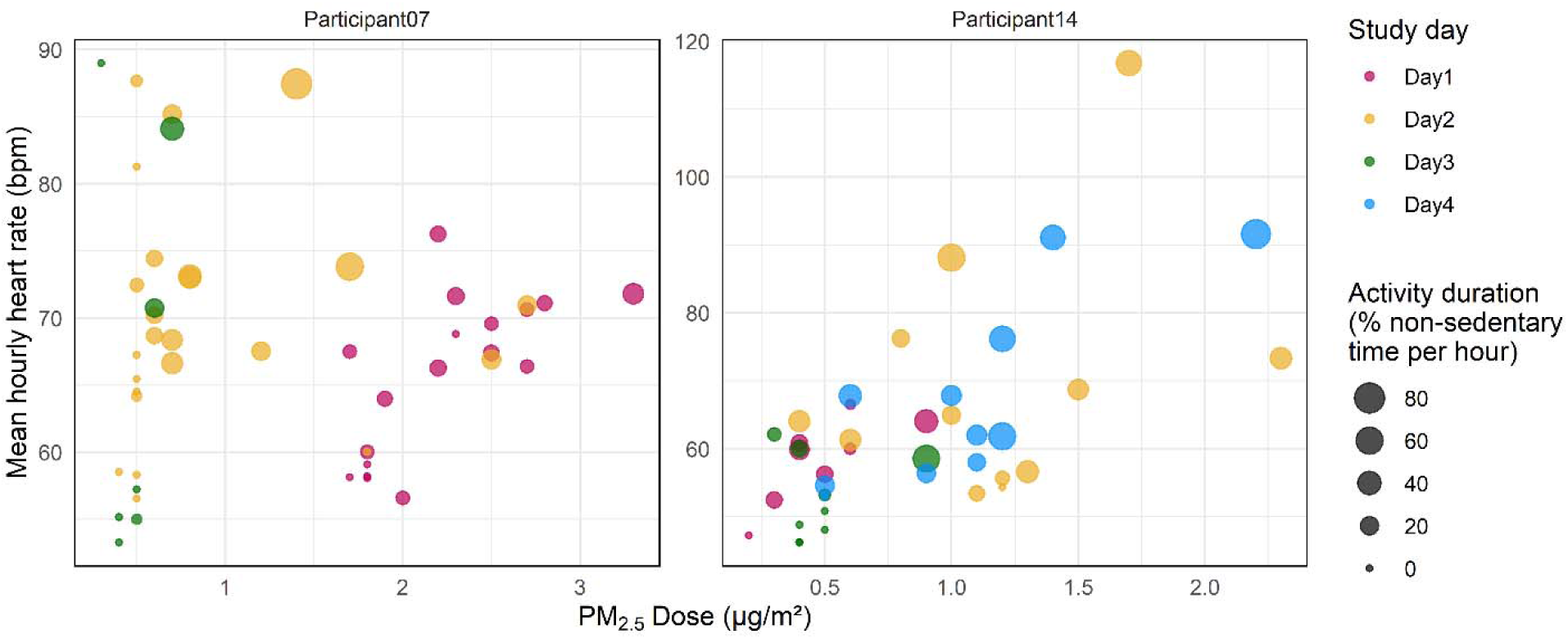
Relationship between hourly inhaled doses of PM_2.5_, heart rate & physical activity. Hourly inhaled doses of PM_2.5_ plotted against mean hourly hear rate where size of each data point indicates activity duration calculated as % non-sedentary time per hour. Participant #14 shows the expected relationship between the three variables based on the fact that hourly inhaled doses of PM_2.5_ were calculated by using heart rate as a proxy to estimate physical activity. However, Participant #7 displays high degree of inter-day variability. Data panels for the remainder of study participants are shown in **Error! Reference source not found.**

### Diurnal variability in exposure metrics

Given the observations that concentrations of particulate matter and ozone show diurnal patterns in urban settings ^47^, we set out to parse diurnal variability in our dataset. As expected, heart rate followed an oscillatory pattern in our cohort (**Error! Reference source not found.**) where the day mean (±SD) of 74.6±18.7 bpm dropped at night to 61.2±10.7 bpm.

Hourly ozone concentrations displayed diurnal patterns (as expected for ground-level ozone due to the photochemical reaction ^48^ across the cohort in the outdoor levels (Tier 1, day 34.4±12.6 ppb, night 24.3±10.5 ppb), exposure (Tier 7, day 9.3±10.7 ppb, night 5.7±6.8 ppb) and inhaled dose (Tier 8, day 6.8±8.6 µg/m², night 3.2±4.5 µg/m²) as visualized in *Error! Reference source not found.* and *Error! Reference source not found..* For PM_2.5_, diurnal patterns across the cohort were small as evident for outdoor levels (Tier 1, day 8.1±5.3 µg/m³, night 8.4±4.7 µg/m³), exposure (Tier 7, day 5.5±4.0 µg/m³, night 5.4±3.4 µg/m³) and the inhaled dose (Tier 8, day 2.1±1.8 µg/m², night 1.5±1.1 µg/m²).

### Between-subject variance in PM_2.5_ and ozone outdoor exposure independent of neighborhood socioeconomic status

The continuous collection of GPS coordinates allowed us to capture the broader socioeconomic context which participants were exposed to during the study period. We refer to this as neighborhood socioeconomic status (Error! Reference source not found.). To explore whether the socioeconomic status was a major determinant of the high degree of inter-individual variability observed in PM_2.5_ and ozone exposure and dose, we correlated outdoor concentration levels with the census-block level area deprivation index (ADI), provided by The Neighborhood Atlas ^40^, as a proxy for individual-level social environment. To assess this, we assigned ADI percentiles to each measurement using the GPS coordinates of the AirNow, OpenAQ and PurpleAir stations the PM_2.5_ and ozone readings. Notably, directional relationships between national ADI percentiles, and outdoor levels of PM_2.5_ and ozone were not discernable for the AirNow, OpenAQ and PurpleAIr networks (**Error! Reference source not found.**).

PM_2.5_ exposure levels were, for example, comparable for the 20^th^ and 70^th^ national ADI percentile during the day (22.1±10.0 µg/m^3^ and 24.4±7.3 µg/m^3^, respectively). Notably, a majority of PurpleAir sensors were assigned to a valid ADI (96%, 23 out of 24) compared to fewer cases for the AirNow sensors (82%, 14 out of 17,**Error! Reference source not found.**).

### Feasibility for community-driven outdoor monitoring of PM_2.5_ levels

The correlation of hourly dose estimates of PM_2.5_ outdoor levels showed substantial agreement between AirNow and PurpleAir derived modeling (Pearson’s correlation, *R*=0.84, *p*<2.2e-16, **Error! Reference source not found.*a left***) where neither participant, region nor time-of-day had an outsized influence as grouping variable (**Error! Reference source not found.*a right***). While measurements of PM_2.5_ levels were recorded by a similar number of AirNow deployed stations and community-embedded PurpleAir sensors (paired t-test *p*=0.74), the mean distance between study participant and PurpleAir sensor was less compared to the AirNow stations (paired t-test *p*=0.007, **Error! Reference source not found.*b***). This offers the following considerations: i) a community-driven approach as a scalable, affordable solution to supplement existing air monitoring networks, ii) potentially improving accuracy of inhaled dose predictions for densely populated urban and metropolitan areas, and iii) extending air monitoring networks more easily into rural communities.

## Discussion

In this exploratory study, we demonstrate the feasibility of deploying TracMyAir, a smartphone-based digital health platform, to generate temporally resolved, individual-level exposure and inhaled dose estimates for PM_2.5_ and ozone under real-world conditions. Across 18 participants and more than 1,500 participant-hours, TracMyAir successfully integrated geolocation, ambient air quality data, building infiltration modeling, microenvironment classification, and wearable-derived physical activity to produce eight tiers of hourly exposure metrics, ranging from outdoor concentrations to personalized inhaled doses. To provide more fine-scale exposure metrics, TracMyAir accounted for spatial variations in ambient PM_2.5_ and ozone concentrations by using the smartphone location history to determine the nearest outdoor PM_2.5_ and ozone monitors each hour.

The TracMyAir estimates demonstrated substantial inter-individual and intra-individual variability in both the hourly exposure to and inhaled doses of PM_2.5_ and ozone, personalized to temporal variations in physical activity, individual home conditions, and other factors such as time spent in different microenvironments and meteorological conditions. In our dataset, exposure and dose tracked closely but were not synonymous. Their partial divergence indicates that dose-based metrics may capture subtle fluctuations in PM_2.5_ and ozone burden that concentration-based exposure alone can miss. While exposures (Tier 7) reflected the local pollutant levels in the microenvironments the participants occupied (including the strong indoor attenuation of ozone relative to outdoors), inhaled dose (Tier 8) also incorporated physiological demand and activity through proxy measurements (step counts or heart rates). Consistent with this distinction between data tiers, modeled doses largely agreed with exposures but only partially explained observed variances in the data (e.g., for PM_2.5_, ∼51% variance explained between exposure and dosage using heart rate and ∼73% using phone-or watch-based step counts; for ozone, ∼68% and ∼84%, respectively), indicating that incorporating physical activity meaningfully changes the inferred pollutant burden relative to concentration-only exposure estimates.

The importance of this distinction is supported by prior work demonstrating that lung function tracks more closely with personal-level exposure metrics than with ambient central-site concentrations. In a study of children with asthma ^49^, Delfino et al. showed that personal exposures to particulate air pollution were significantly associated with reduced lung function, whereas corresponding ambient concentrations measured at central monitoring sites were weaker predictors or not significantly associated with pulmonary outcomes. These findings highlight a key limitation of relying on ambient concentrations as proxies for individual exposure and reinforce the need for exposure assessment approaches that resolve where individuals are located, what microenvironments they occupy, how those environments modify PM_2.5_ and ozone levels, and their levels of physical activity.

Inhaled dose reflects not only the concentration in the surrounding air but also the body’s moment-to-moment ventilatory demand. This distinction is especially consequential for respiratory endpoints, where the effective delivery of PM_2.5_ and ozone to the airways is shaped by physical activity and by diurnal physiology including circadian variation in airway caliber, autonomic tone, and breathing patterns ^4,20–22,25–30^. However, the incremental information provided from personalized inhaled dose estimates appeared modest in this cohort. Determining whether TracMyAir’s inhaled dose estimates meaningfully enhance the prediction of functional or clinical health outcomes will require further validation in mechanistic studies under real-world conditions.

We observed that inhaled dose estimates derived from Apple Watch heart rate were higher on average and more variable than those derived from iPhone or Apple Watch step counts. This likely reflects the fact that heart rate is sensitive not only to physical activity but also to non-ambulatory stressors ^50,51^, for example, driving in heavy traffic can elevate heart rate via sympathetic activation even when step counts are negligible, thereby potentially increasing inferred ventilation and dose relative to step-based approaches.

Over the past two decades, personal exposure assessment has advanced substantially in both form factor and scalability. What once required 3.211kg backpack-based monitoring systems in controlled field studies ^52^ can now be accomplished with smartphone-centered approaches ^53^, enabling high-resolution spatiotemporal exposure characterization at a scale that is far more practical for real-world epidemiologic and health outcome research.

Our comparison of AirNow and community-embedded PurpleAir data further supports the growing recognition that dense, low-cost sensor networks can meaningfully augment regulatory monitoring, particularly in urban environments where spatial gradients are steep. The shorter average distance between participants and PurpleAir sensors, combined with the high correlation in modeled doses across data sources, suggests that community-driven monitoring can enhance spatial representativeness without sacrificing consistency. TracMyAir’s current architecture treats AirNow and PurpleAir as separate inputs, but future integration of both data streams within a unified modeling framework could further improve robustness and resilience to data gaps.

The large inter-individual differences in PM_2.5_ and ozone levels raised the question to what extent these differences were explained by the socioeconomic context of participants’ neighborhoods. Surprisingly, we found no discernable directional relationship between Area Deprivation Index (ADI) percentiles and outdoor PM_2.5_ or ozone levels. This suggests that in this cohort and study setting, other determinants, such as regional meteorology, temporal dynamics, mobility, and microenvironmental factors, may have been the predominant sources of variability captured by TracMyAir.

This study has several limitations. First, the sample size was modest and consisted of apparently healthy, non-smoking adults, limiting generalizability to vulnerable populations. Second, while the study design included deployment of indoor and outdoor PurpleAir sensors at participant homes, logistical constraints meant that full indoor-outdoor paired monitoring was achieved in only a small subset of participants. Although modeled indoor concentrations compared favorably with available measurements, broader validation across diverse housing types remains necessary. Third, TracMyAir currently operates only on the iOS ecosystem. While iPhones and Apple Watches are widely used, this platform dependence may introduce selection bias and limits applicability in populations with lower smartphone access or different operating system preferences. Extending TracMyAir to additional operating systems represents an important step toward equitable and scalable deployment. To reduce this selection bias for the present study, we provided deployment-ready study-iPhones to our participants. Fourth, although TracMyAir dynamically selects the nearest monitoring stations based on participant geolocation, we did not explicitly quantify how exposure estimates vary as a function of distance to monitors. Finally, this study was conducted during the COVID-19 pandemic, when time-activity patterns were atypical, with participants spending more time indoors compared to historical norms. While this context underscores the importance of indoor environments in exposure assessment, it may limit extrapolation to non-pandemic conditions.

In conclusion, we demonstrated under real-world conditions that TracMyAir consistently models both exposure and inhaled dose estimates for PM_2.5_ and ozone, dynamically accounting for the diverse microenvironments individuals traverse throughout the day. The observed divergence between concentration-based exposure metrics and inhaled dose-based estimates underscores the platform’s ability to capture subtle, hyperlocal fluctuations in the personal PM_2.5_ and ozone milieu.

Future follow-up studies that systematically compare personal exposure estimates against monitors at varying distances and ideally against direct personal measurements will be critical to test rigorously the added value of hyperlocal modeling. At a larger scale, downloading and analyzing AirNow and PurpleAir data across broader regions could allow formal assessment of spatial variability and decay with distance, strengthening the case that hyperlocal approaches yield more accurate and health-relevant exposure estimates.

## Data Availability

The data used for the analysis are accessible at www.tbd.org.

## Code Availability

The code used for the analysis is accessible at www.tbd.org.

## Data Availability

All data produced in the present study are available upon reasonable request to the authors

## Acknowledgments

We thank the participants for volunteering. Caroline Chivily successfully fulfilled the required fieldwork practice experiences through this study as part of her Master of Public Health (MPH) program at the University of Pennsylvania. LaVenia Banas provided excellent logistical support. C.S. is the Robert L McNeil Jr. Fellow in Translational Medicine and Therapeutics. G.A.F. is the Robert L McNeil Jr. Professor in Translational Medicine and Therapeutics.

## Author Contributions

CS designed the study and obtained ethics approval. VI provided guidance and expert knowledge on adopting the TracMyAir platform. CC and NEJ recruited study participants with support from CS. NFL, VI and CS analyzed the data and interpreted the results with bioinformatics support from AM. NFL, VI and CS wrote the manuscript. NFL and VI prepared figures and tables. All authors, i.e. NFL, VI, CC, NEJ, AM, GAF and CS reviewed the manuscript and provided feedback which was the basis for several rounds of revisions by NEJ, VI and CS prior to submission.

## Disclaimer

The views expressed in this article are those of the author(s) and do not necessarily represent the views or policies of the U.S. Environmental Protection Agency. Mentioning trade names or commercial products does not constitute endorsement or recommendation for use.

