## Supplemental Materials for "TracMyAir: Smartphone-enabled spatiotemporal estimates for inhaled doses of particulate matter and ozone to personalize health outcomes"

for

### ***Supplemental Results***

#### ***Modeling time-specific exposure metrics of PM2.5 and ozone using geolocation and microenvironment (Tiers 1-7)***

The TracMyAir modeling based on data from the AirNow network predicted mean (±SD) distributions of hourly $PM_{2.5}$ and ozone outdoor concentrations of 8.2±5.2 μg/m³ and 31.8±13.1 ppb, respectively. Estimates of $PM_{2.5}$ based on the PurpleAir network were comparable at 7.5±4.8 μg/m³. The subject-level estimates for mean (±SD) hourly outdoor $PM_{2.5}$ and ozone levels (Tier 1) ranged from a minimum of 3.9±1.9 μg/m³ to a maximum of 12.3±9.4 μg/m³ and from a minimum of 24.9±7.2 ppb to a maximum of 39.2±13.5 ppb, respectively, highlighting the large between-subject variability (***Supplementary Figure 16***). Similar large inter-individual variability was observed for the hourly home infiltration factor (Tier 3, mean±SD ranging from 0.4±0.2 to 0.83±0.01 and from 0.05±0.01 to 0.75±0.13 for $PM_{2.5}$ and ozone, respectively), home indoor microenvironment (Tier 4, ranging from 2.0±1.8 μg/m³ to 9.6±2.6 μg/m³ and from 1.3±0.4 ppb to 28.3±10.2 ppb for $PM_{2.5}$ and ozone, respectively) and exposure (Tier 7, ranging from 2.5±1.2 μg/m³ to 9.5±2.5 μg/m³ and from 1.3±0.4 ppb to 31.9±14.2 ppb for $PM_{2.5}$ and ozone, respectively) (***Supplementary Figure 16***). Notably, the hourly infiltration factors vary considerably both among participants and their individual homes. This variability is likely attributable to temporal variations in indoor-to-outdoor temperature differences, wind speeds, infiltration (e.g., building characteristics) and natural ventilation (e.g., open windows). For context, each home's hourly indoor concentrations (Tier 4) were lower than the outdoor levels (Tier 1) for both $PM_{2.5}$ and ozone since they had home infiltration factors (Finf_home) < 1. This difference between home indoor and outdoor concentrations was smaller for $PM_{2.5}$ than ozone because the home infiltration factor was higher for $PM_{2.5}$ than ozone in line with previous reports [1].

In addition to air quality observations from AirNow, TracMyAir retrieved data from community-based PurpleAir monitors close to each participant's microenvironment. ***Supplementary Figure 17*** shows similarly wide-ranging distributions of hourly $PM_{2.5}$ concentrations for each study participant, ranging for outdoor concentrations in Tier 1 from 1.2±0.8 μg/m³ to 11.2±6.7 μg/m³, for home indoor microenvironment in Tier 4 from

0.7±0.4 μg/m$^3$ to 8.7±2.7 μg/m$^3$, and for exposure in Tier 7 from 0.8±0.5 μg/m$^3$ to 8.7±2.7 μg/m$^3$ $PM_{2.5}$. Similar to AirNow-based indoor concentration estimates, each home's estimated indoor concentration was lower than outdoor for $PM_{2.5}$. The comparison of hourly indoor $PM_{2.5}$ concentrations between data collections from study-deployed (n=3 participants' homes) versus community-embedded PurpleAir sensors showed good agreement ($R^2$=0.91, $p$=0.003; $R^2$=0.76, $p$=2.2e-16; $R^2$=0.57, $p$=1.4e-8; ***Supplementary Figure 18***).

The higher tiers of exposure metrics (Tier 5: personal exposure factor, Tier 6: time spent in microenvironments, Tier 7: exposure) account for time spent in different indoor and outdoor locations. The hourly fraction of time spent in each microenvironment was determined from smartphone geolocations, speed, and building boundaries using the microenvironment classification model called MicroTrac [2]. In the Indoor versus Outdoor comparison (***Table S 7***), subjects spent an average of 91.7% of measurements indoors, 5.8% outdoors, 0.6% in vehicles and 1.9% in mixed microenvironments based on the AirNow estimates (note that averages across all participants do not necessarily add up to 100%). The percentages were comparable for the PurpleAir based estimates. In the Home versus Work comparison, participants stayed "At Home" for an average of 67.2% of measurements, and 19.7% "At Work". Using PurpleAir, "At Work" was estimated higher at 25.1% of measurements (***Table S 7***). Likely due to the COVID19 pandemic, participants spent overall more time indoors (despite enrollment over the summer) compared to survey data in the 1990s where an average of 87% of respondents' time was spent in enclosed buildings [3].

Cohort distributions of modeled exposures using AirNow-based data collections for $PM_{2.5}$ and ozone are shown in ***Supplementary Figure 19***. Here, hourly mean (±SD) Tier 1 outdoor concentrations amounted to 8.2±5.2 μg/m$^3$ and 31.8±13.1 ppb, respectively; hourly Tier 4 home indoor concentrations to 5.1±3.6 μg/m$^3$ and 10.4±11.2 ppb, respectively; and hourly Tier 7 exposures to 5.5±3.8 μg/m$^3$ and 8.1±9.7 ppb, respectively. For Tier 7, ***Supplementary Figure 7*** shows exposure concentrations specific to the eight microenvironments determined by the TracMyAir model. Here, mean(±SD) "Outdoor

Home" and "Outdoor Work" measured highest hourly $PM_{2.5}$ exposures (9.3±6.6 μg/m³ and 9.1±6.0 μg/m³, respectively), while "Inside Vehicle" was the lowest (3.1±1.9 μg/m³). For ozone, hourly exposures for "Outdoor Home" and "Outdoor Work" were highest (38.4±11.1 ppb and 24.8±12.0 ppb, respectively) and lowest for "Indoors Unknown" and "Indoors Other" (3.8±1.9 ppb and 4.0±1.6 ppb, respectively).

### ***Supplemental Figures***

#### ***Supplementary Figure 1 – Remote sensing domain***

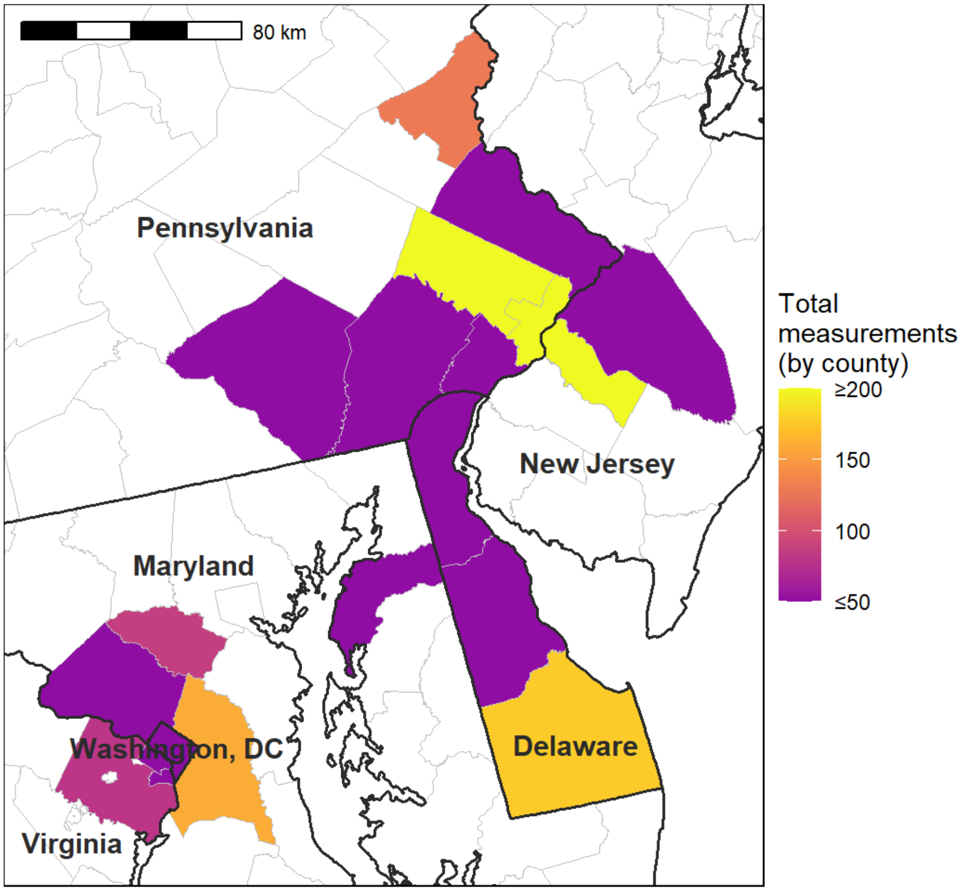

Heatmap indicting the total number of measurements by county accumulated by n=18 participants through geocoded data collections in the TracMyAir mobile app. For reference, the two clusters in the plot comprise Philadelphia, PA, US with its latitude and longitude coordinates of 40.0, -75.2, and Washington DC, DC, US at 38.9, -77.0 (https://www.latlong.net/).

### *Supplementary Figure 2 – Monitoring stations in the remote sensing domain*

a

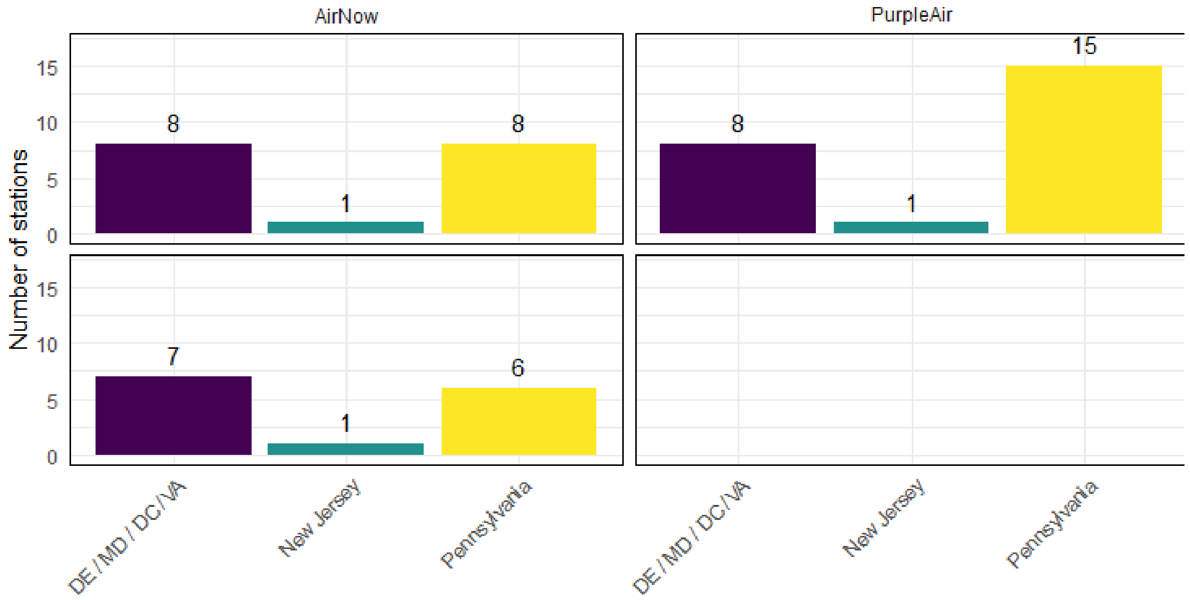

b

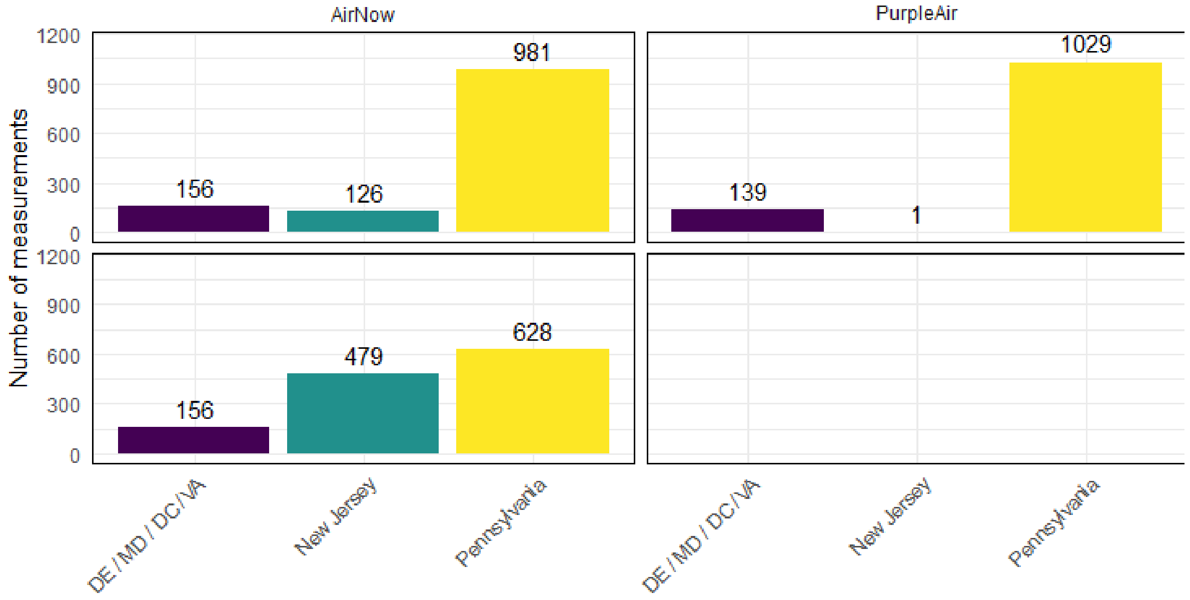

c

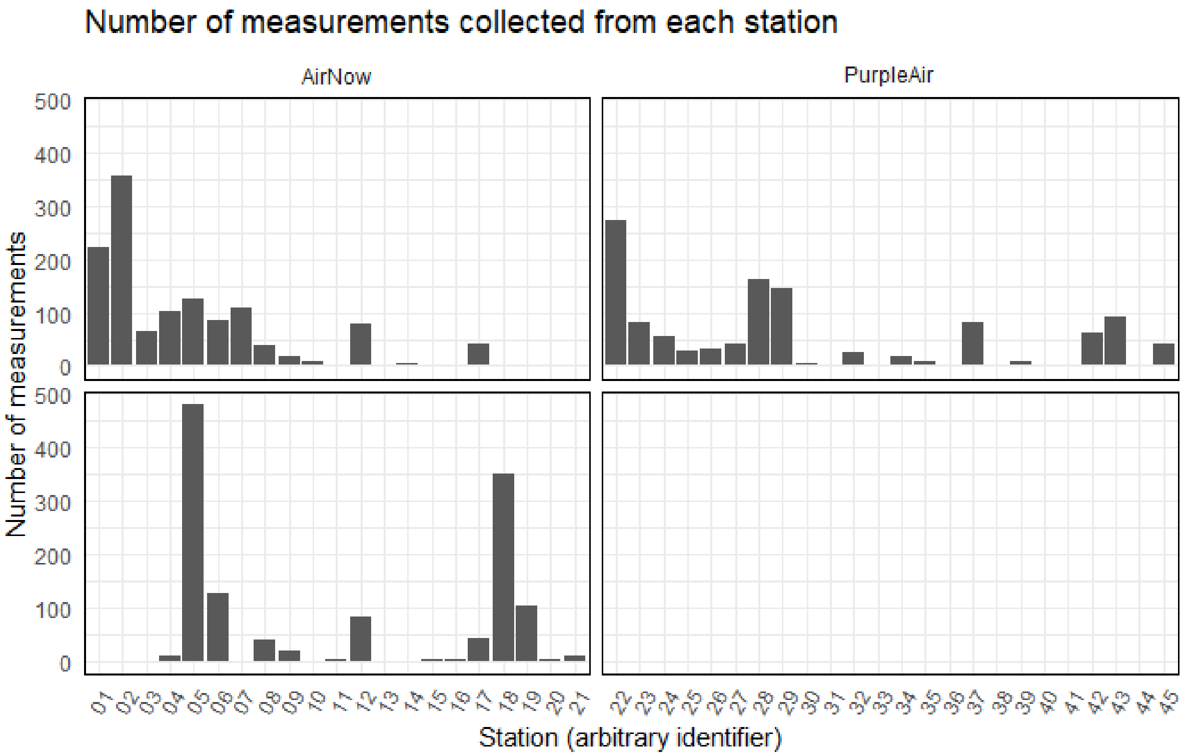

d

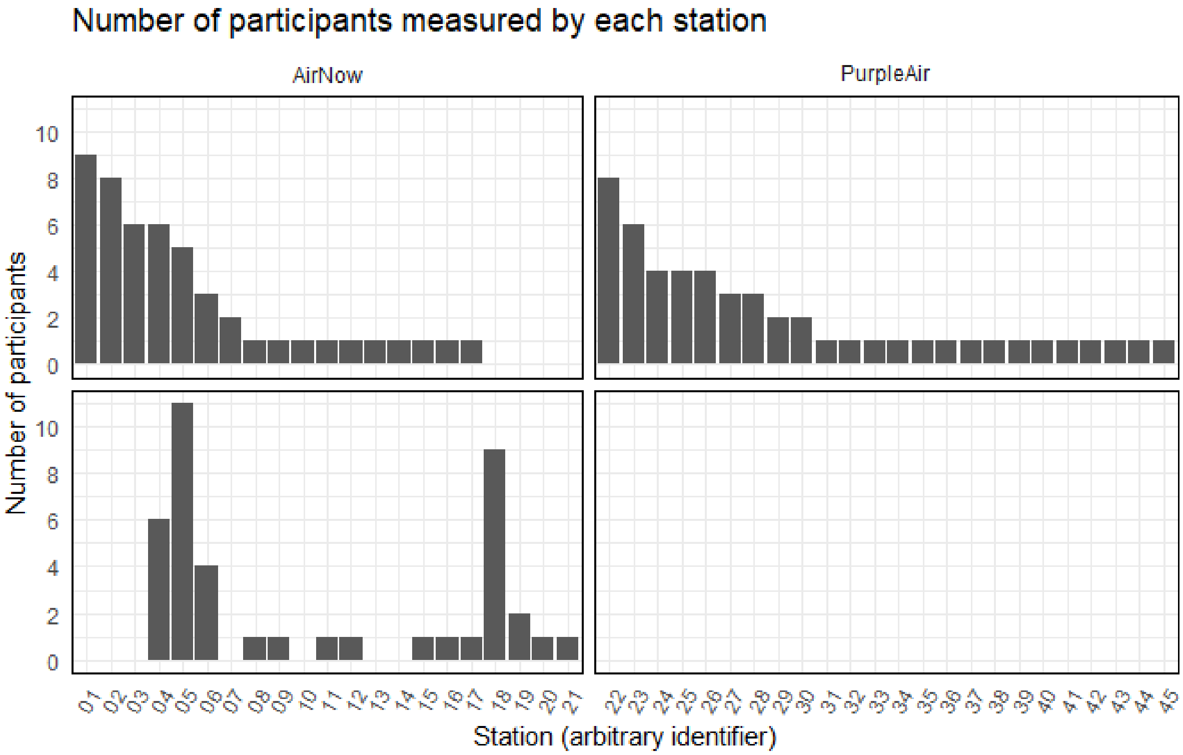

The network of monitoring stations shown for number of stations per state (a), contribution of hourly measurements (b), distributions of measurements collected from each station (c) and participants per station (d).

***Supplementary Figure 3 - Patterns of participant-performed data collection***

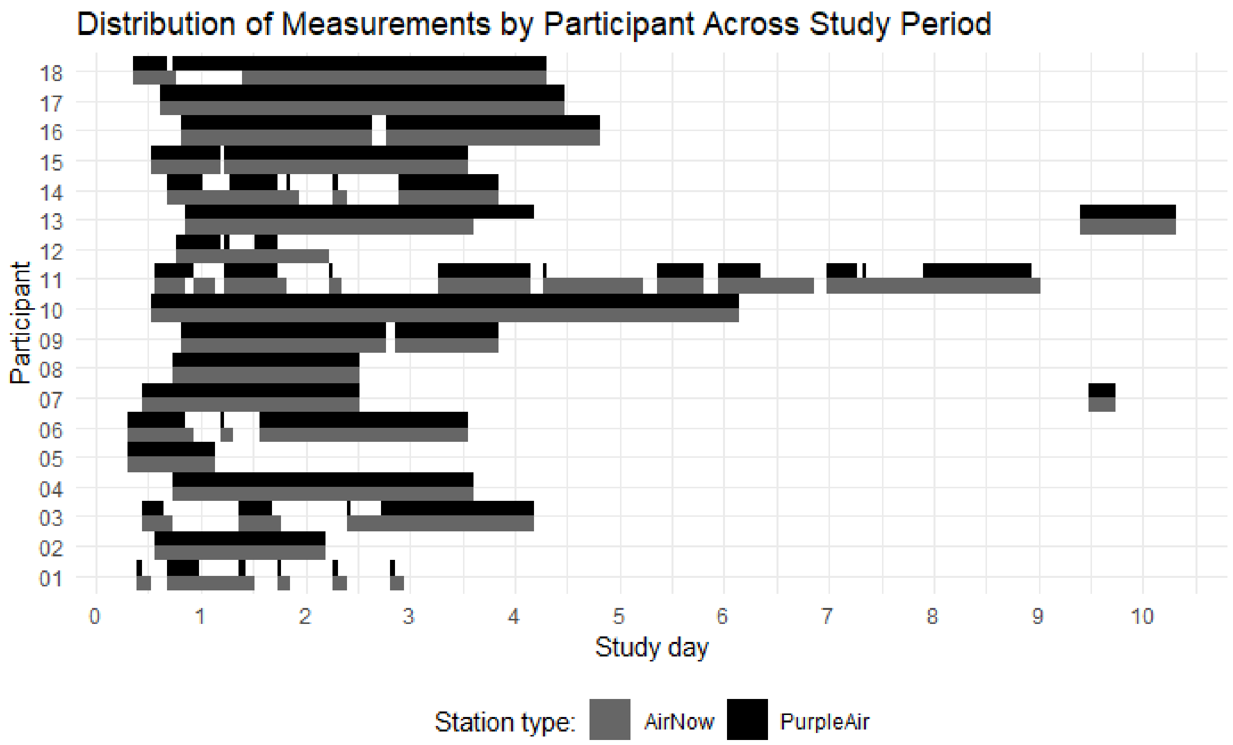

Participant-performed data collection for AirNow monitoring stations and PurpleAir sensors where 0 on the x-axis indicates days since midnight on the first study day.

***Supplementary Figure 4 - Hyperlocal exposure dynamics of $PM_{2.5}$ and ozone***

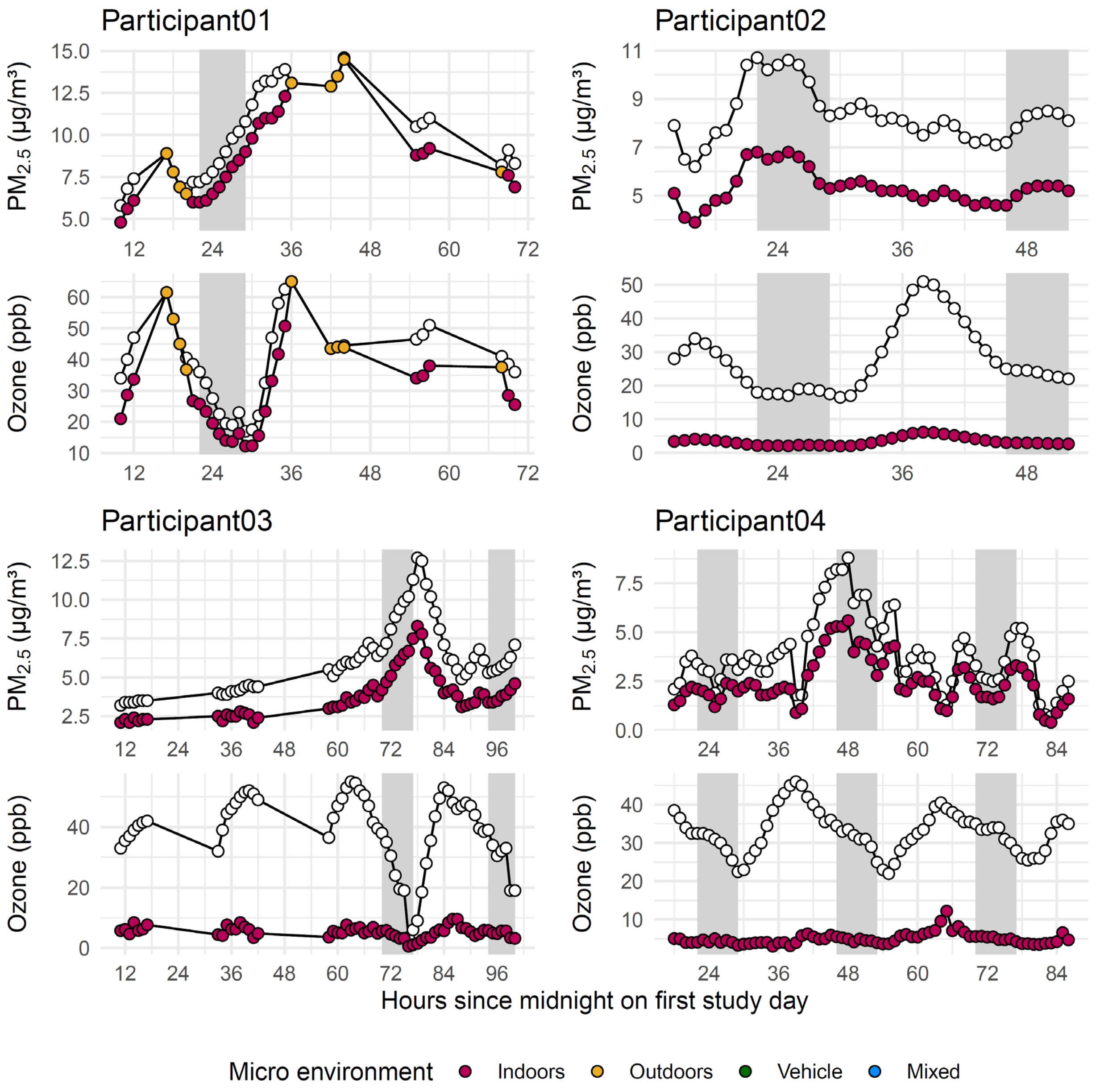

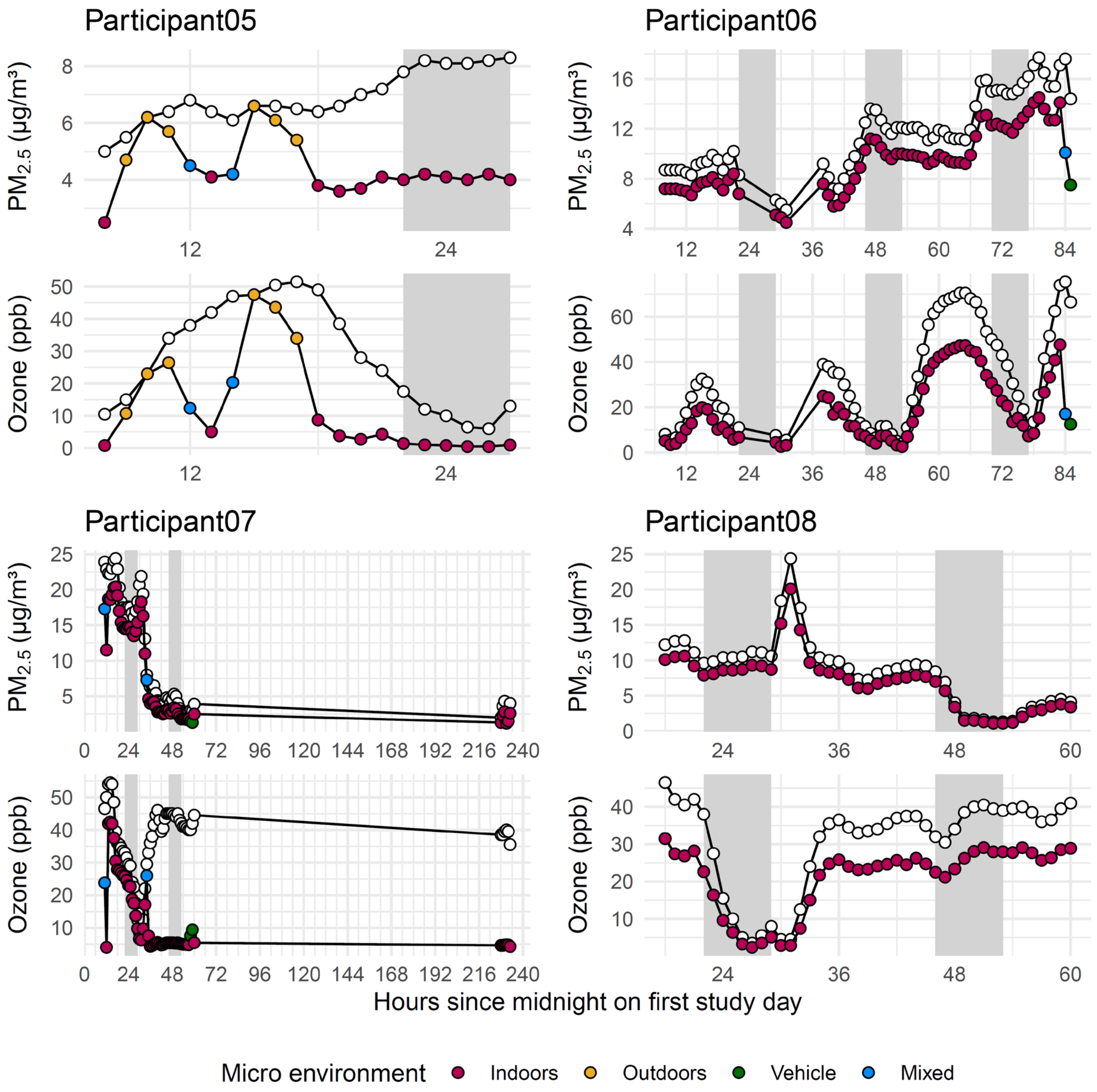
Participant05
Participant06
Participant07
Participant08
PM2.5 (µg/m³)
Ozone (ppb)
Hours since midnight on first study day
Micro environment
Indoors
Outdoors
Vehicle
Mixed

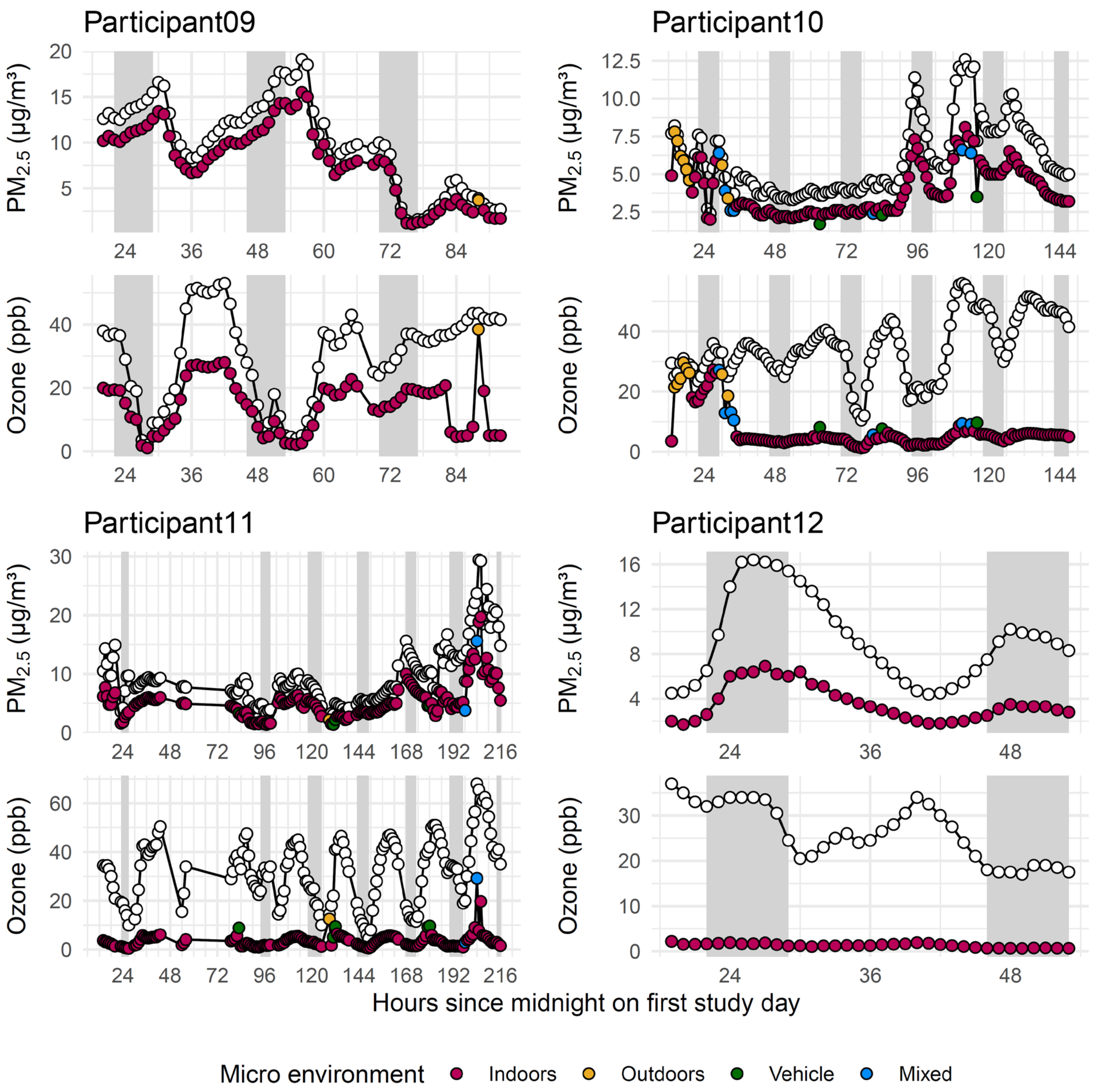

Participant09
Participant10
Participant11
Participant12
PM2.5 (µg/m³)
Ozone (ppb)
Hours since midnight on first study day
Micro environment
Indoors
Outdoors
Vehicle
Mixed

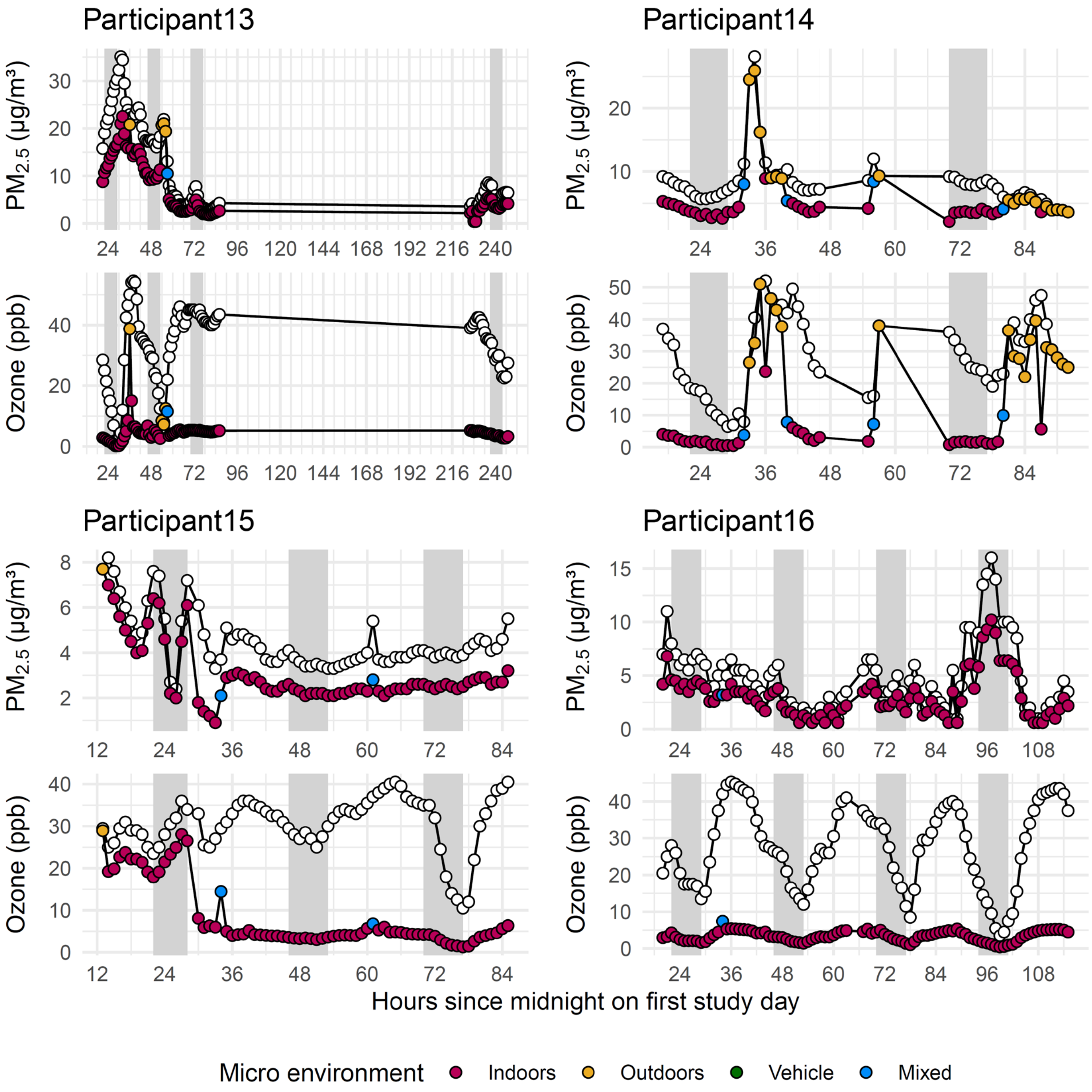
Participant13
Participant14
Participant15
Participant16
PM2.5 (µg/m³)
Ozone (ppb)
Hours since midnight on first study day
Micro environment
Indoors
Outdoors
Vehicle
Mixed

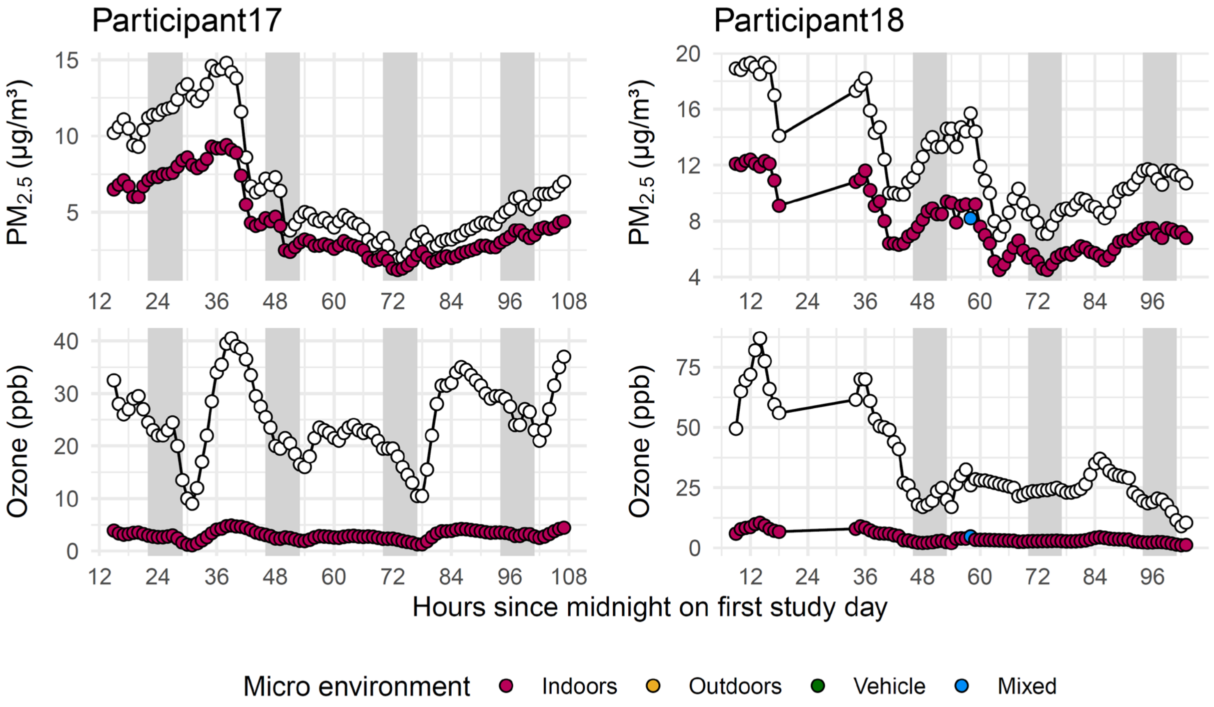

Hyperlocal exposure dynamics of $PM_{2.5}$ and ozone (Tier 7) using AirNow for single participants navigating through indoor (red circle), outdoor (yellow circle), vehicle (green circle) and mixed (blue circle) microenvironment in comparison to raw ambient outdoor levels of $PM_{2.5}$ and ozone (white circles).

***Supplementary Figure 5 – Cohort-level $PM_{2.5}$ and ozone inhaled dose distribution per step count and heart rate***

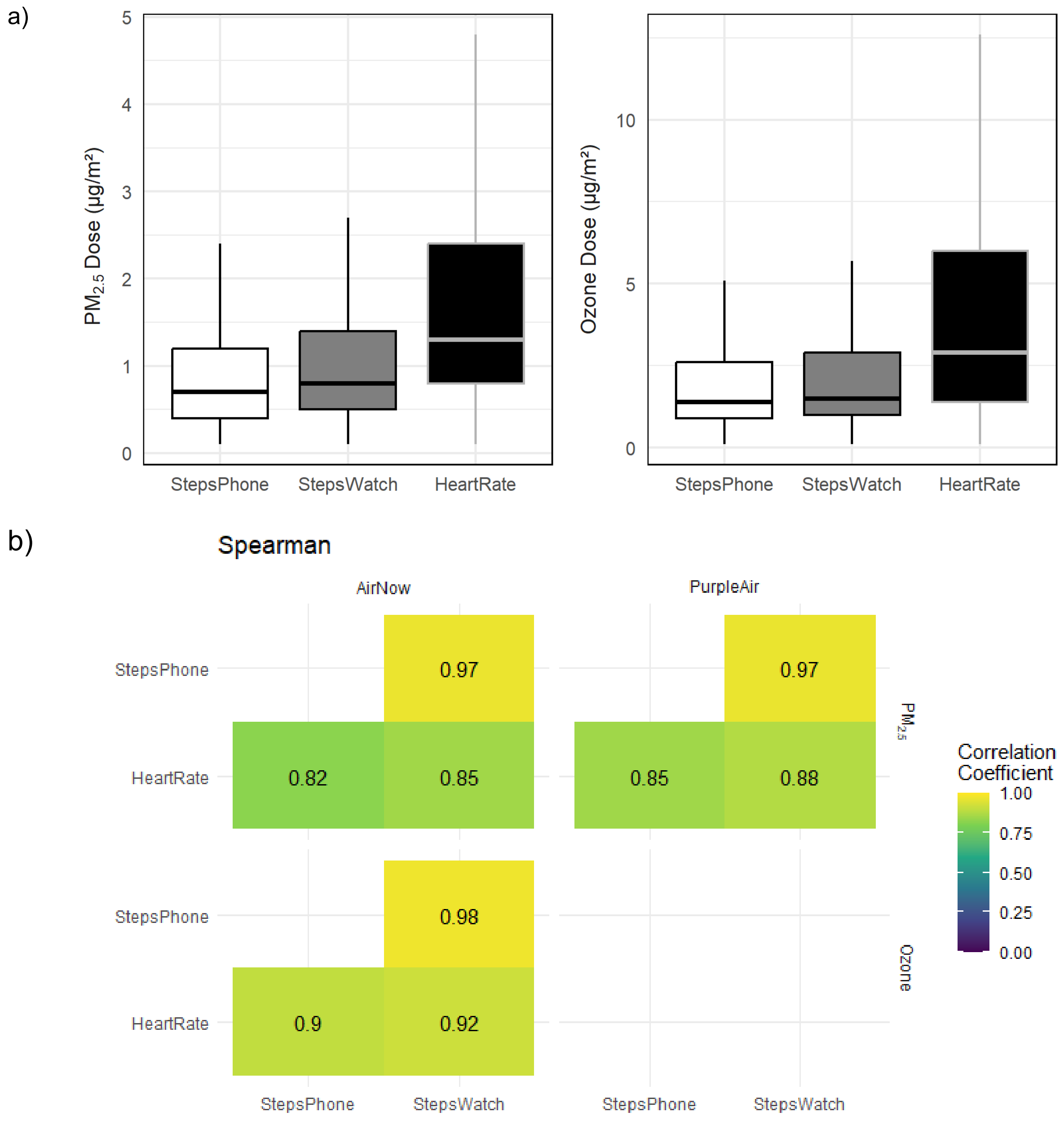

a) Distributions of inhaled dose for $PM_{2.5}$ (left panel) and ozone (right panel) using iPhone step counts (white boxes), Apple watch step counts (gray boxes), and Apple watch heart rate (black boxes). Boxplot lines identify first quartile, median, and third quartile of each distribution. b) Spearman correlation coefficients for estimates of hourly inhaled doses of

$PM_{2.5}$ and ozone comparing step counts from either phone or watch and heart rate as proxies for physical activity using data from the AirNow (left) and PurpleAir (right) sensors.

***Supplementary Figure 6 – Participant-level $PM_{2.5}$ and ozone inhaled dose distribution per step count and heart rate***

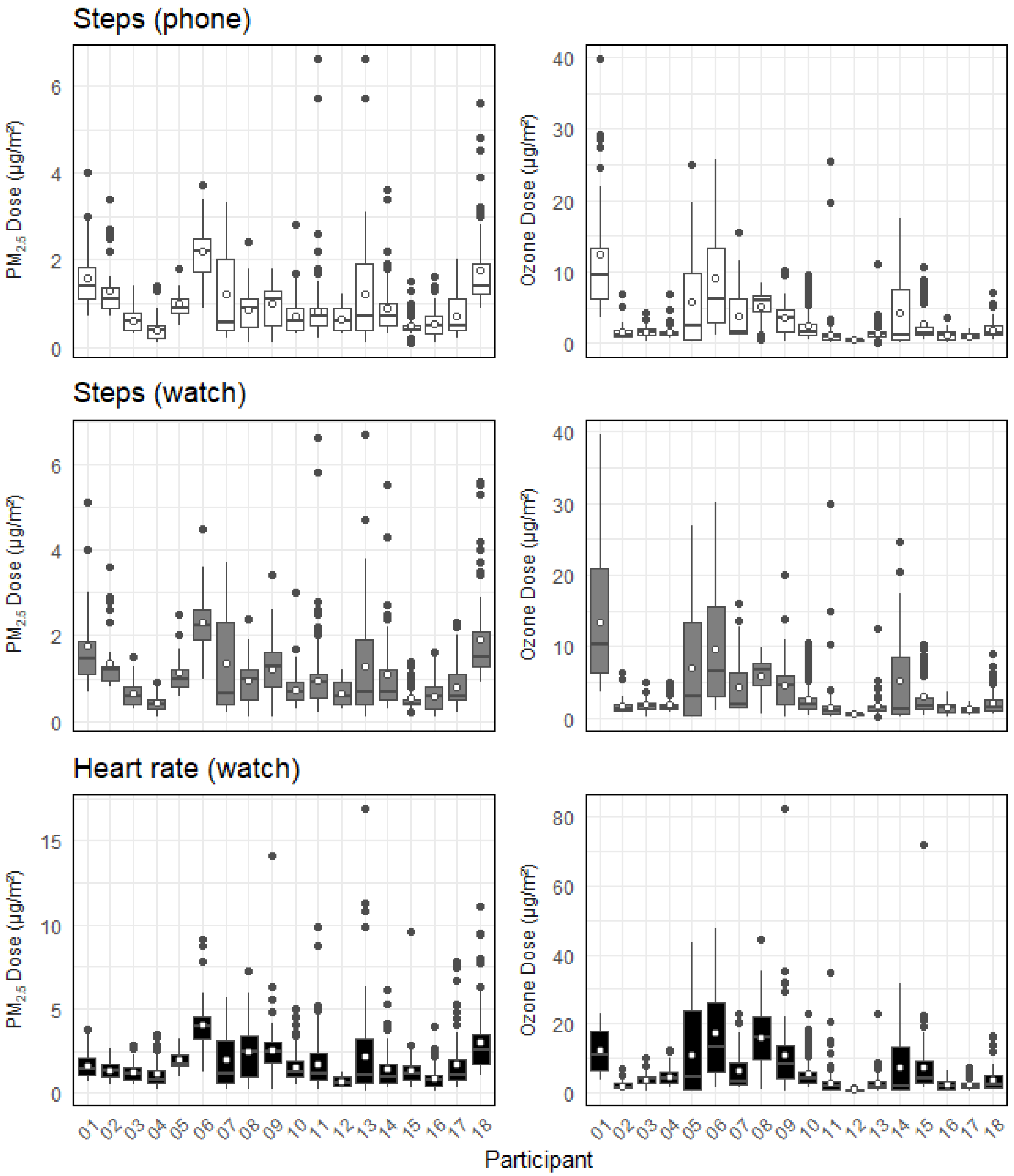

Distributions of inhaled dose for $PM_{2.5}$ (left panels) and ozone (right panels) using iPhone step counts (white boxes), Apple watch step counts (gray boxes), and Apple watch heart rate (black boxes). Boxplot lines identify first quartile, median, and third quartile of each distribution. White dots within boxplots identify distribution means.

***Supplementary Figure 7 – Microenvironment-specific distributions of modeled exposures to PM2.5 and ozone***

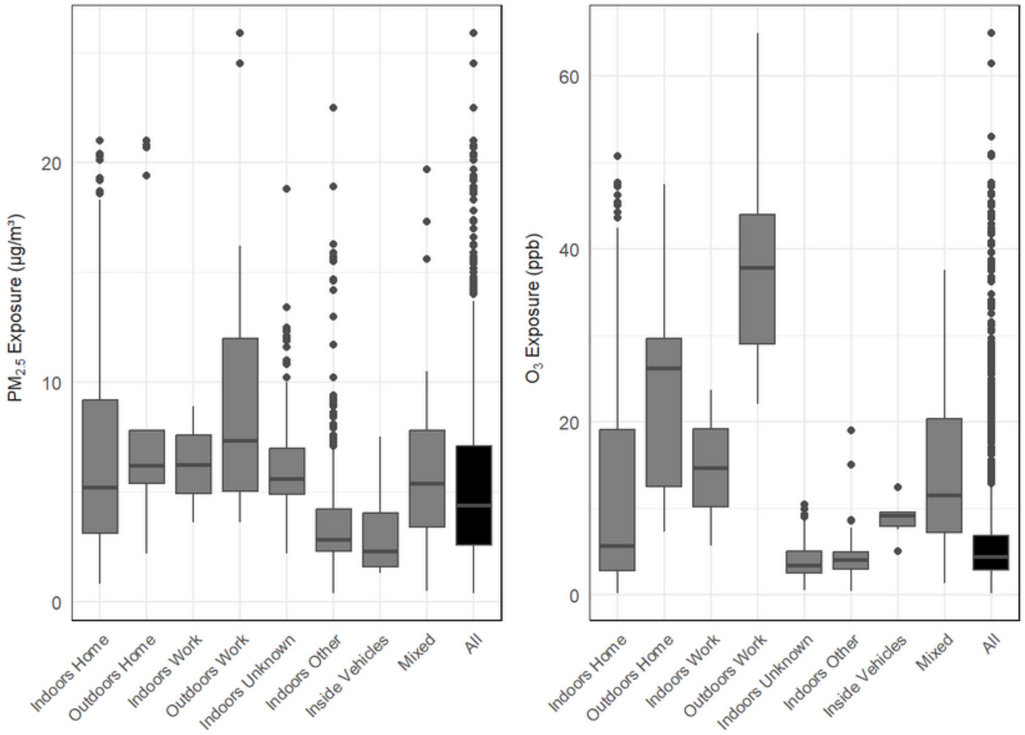

Distributions of $PM_{2.5}$ exposures (left panel) and ozone exposures (right panel) in 6 different Microenvironments (gray boxes), or exposures across all Microenvironments (black box). Boxplot lines identify first quartile, median, and third quartile of each distribution.

#### *Supplementary Figure 8 – Relationship between exposure and hourly inhaled doses of $PM_{2.5}$ and ozone from AirNow*

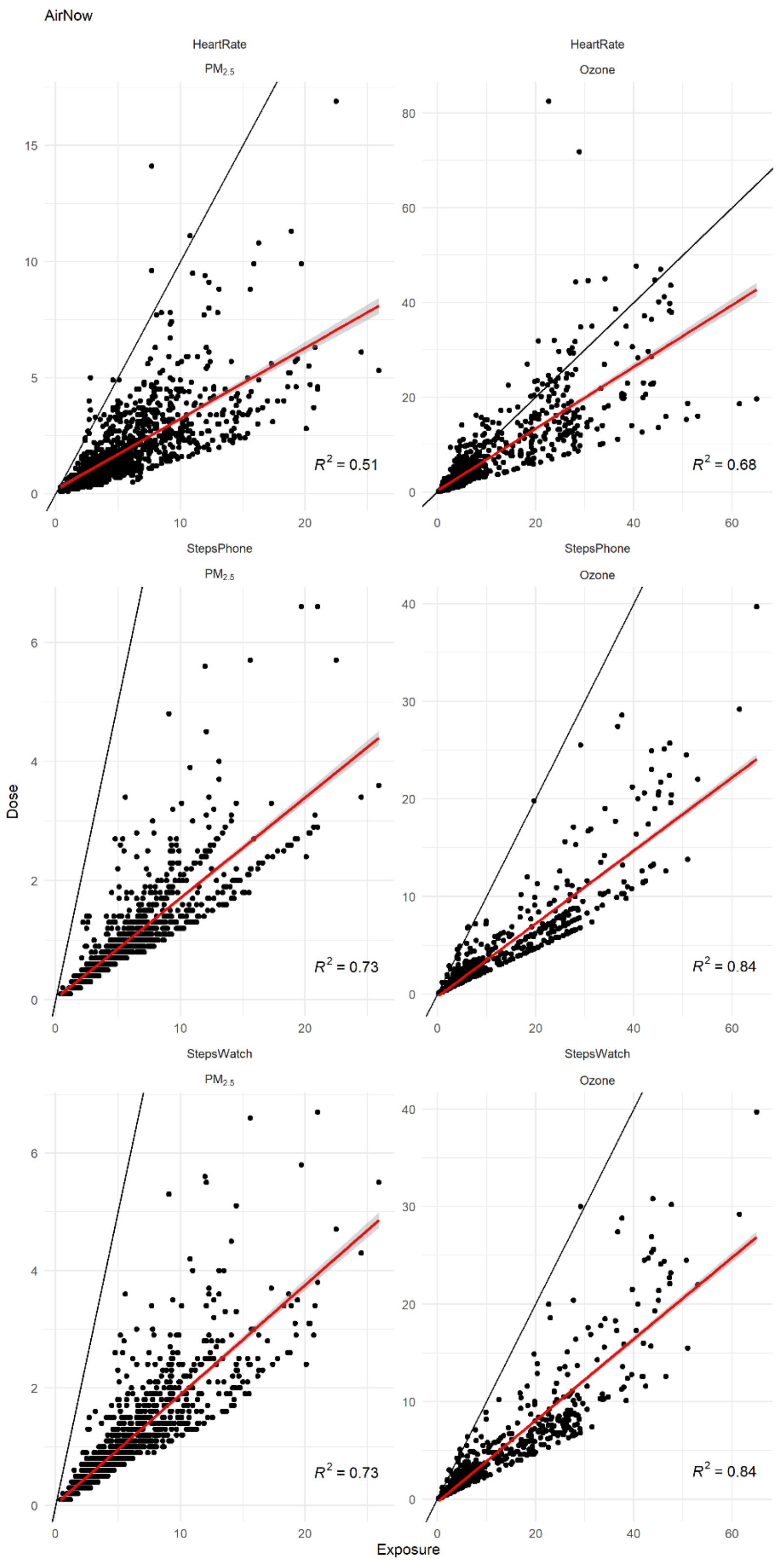

Scatterplots comparing exposures (Tier 7) to inhaled doses (Tier 8) derived from $PM_{2.5}$ (left) and ozone (right) concentrations measured by the AirNow network. Inhaled doses are derived using Apple watch heart rates (top), iPhone step counts (middle), or Apple watch step counts (bottom). Black lines are the diagonals between the two measures, and slim grey areas are the 95% confidence intervals for the regression lines (red). $R^2$ values are coefficients of determination from each simple linear regression.

***Supplementary Figure 9– Relationship between exposure and hourly inhaled doses of $PM_{2.5}$ from PurpleAir***

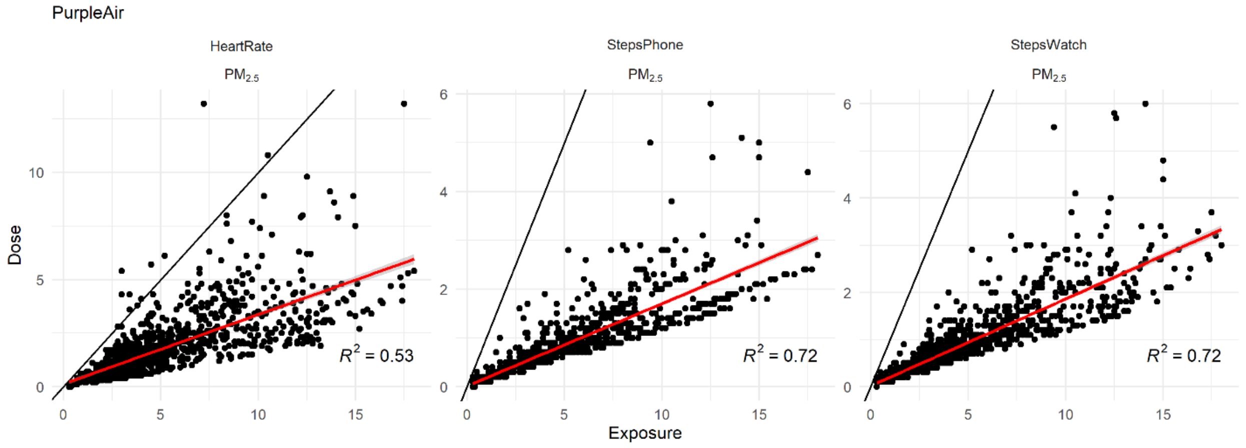

Scatterplots comparing exposures (Tier 7) to inhaled doses (Tier 8) derived from $PM_{2.5}$ concentrations measured by the PurpleAir network. Inhaled doses are derived using Apple watch heart rates (left), iPhone step counts (middle), or Apple watch step counts (right). Black lines are the diagonals between the two measures, and slim grey areas are the 95% confidence intervals for the regression lines (red). $R^2$ values are coefficients of determination from each simple linear regression.

***Supplementary Figure 10 – Relationship between hourly inhaled doses of $PM_{2.5}$, heart rate & physical activity***

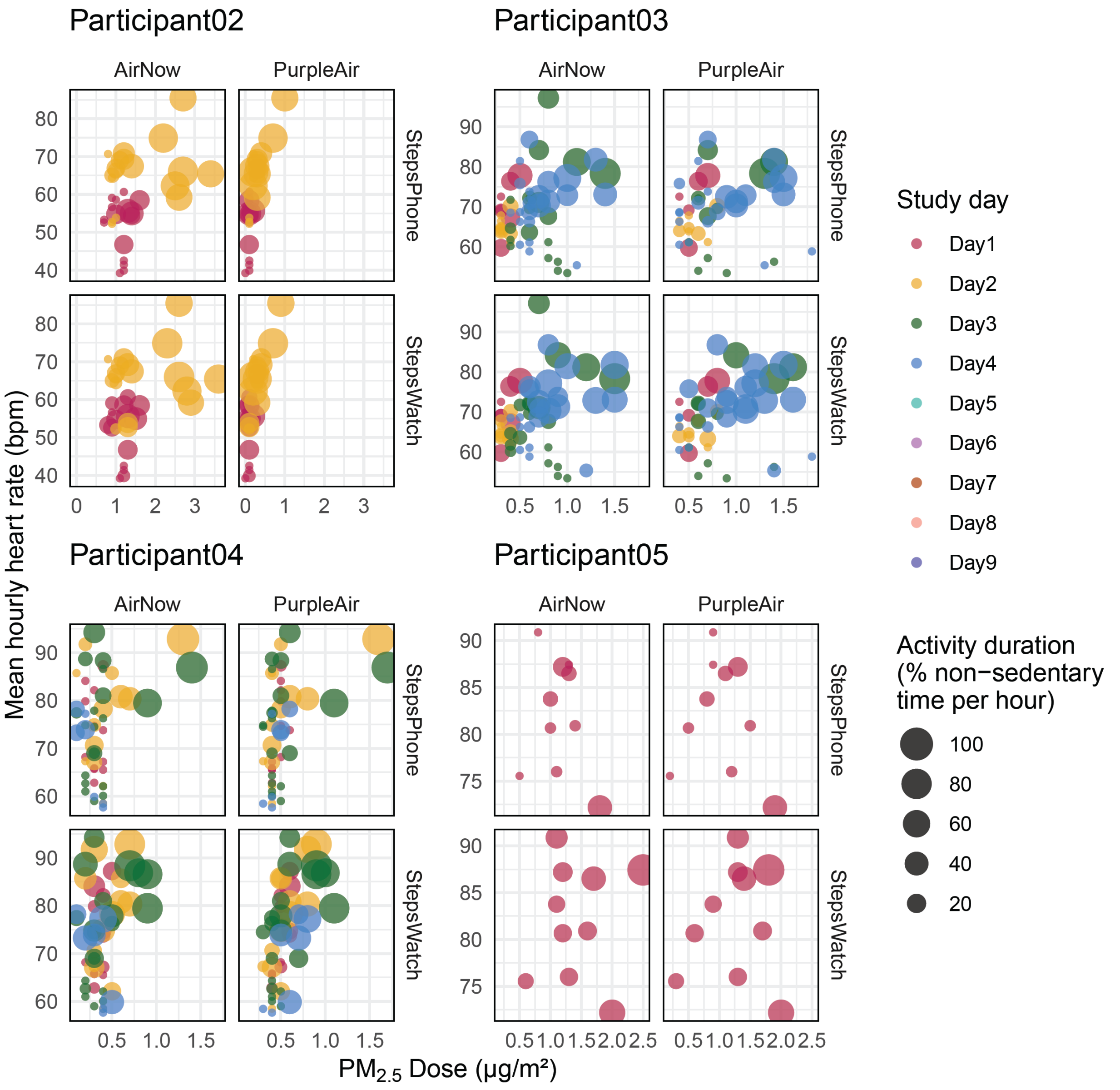

Each dot corresponds to one hour of data. Larger dots indicate higher activity levels

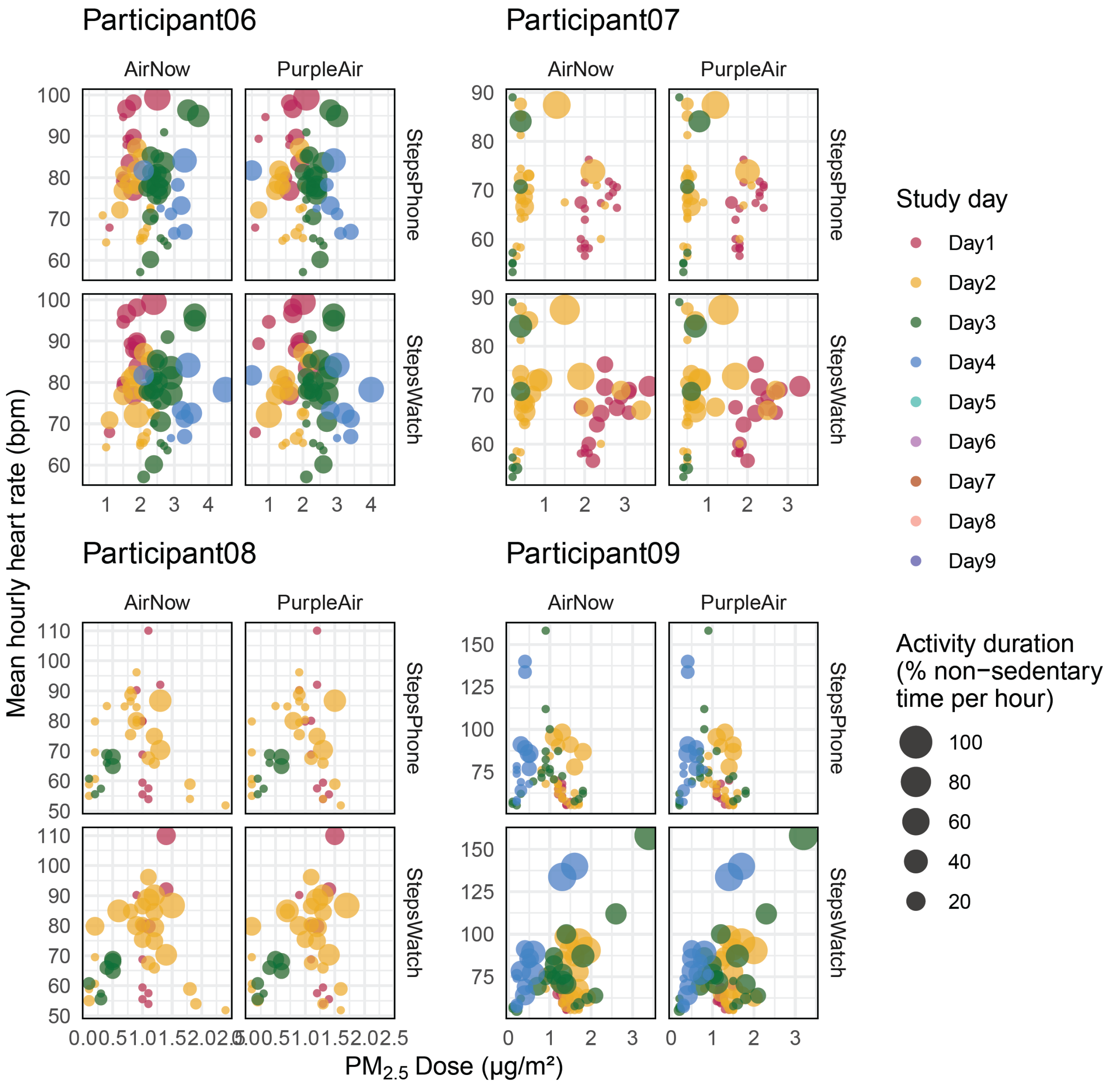

Each dot corresponds to one hour of data. Larger dots indicate higher activity levels

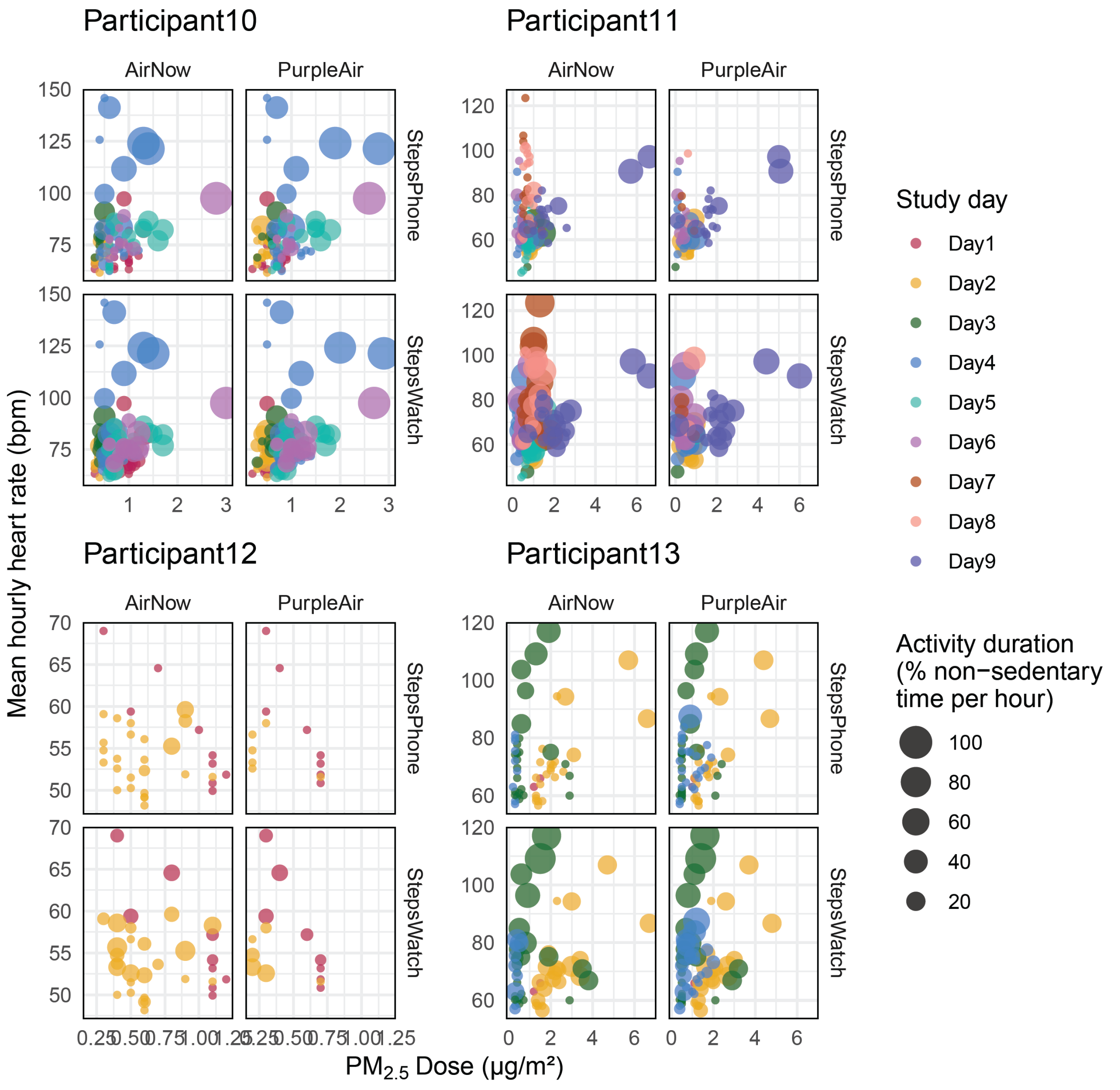

Each dot corresponds to one hour of data. Larger dots indicate higher activity levels

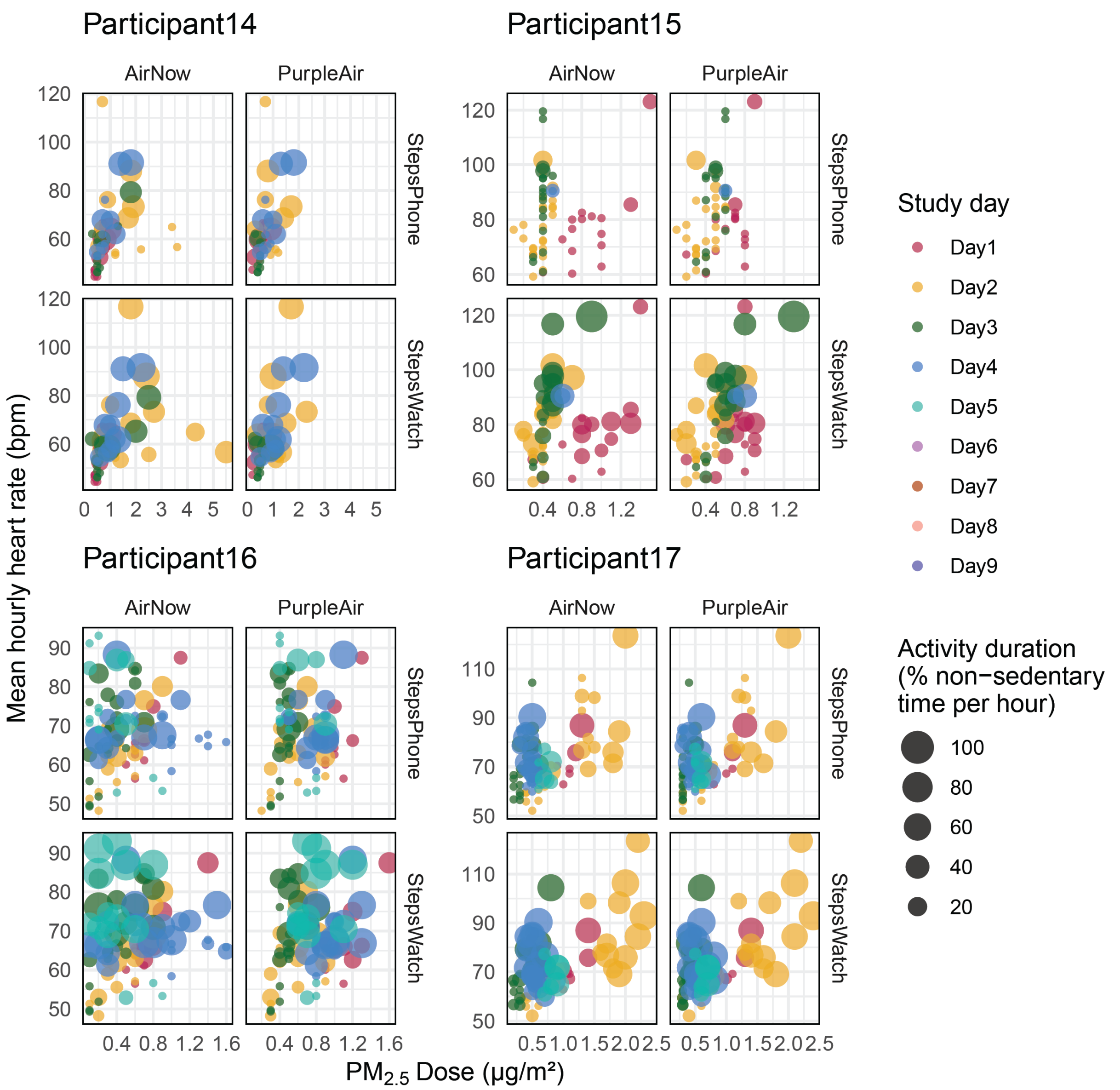

Each dot corresponds to one hour of data. Larger dots indicate higher activity levels

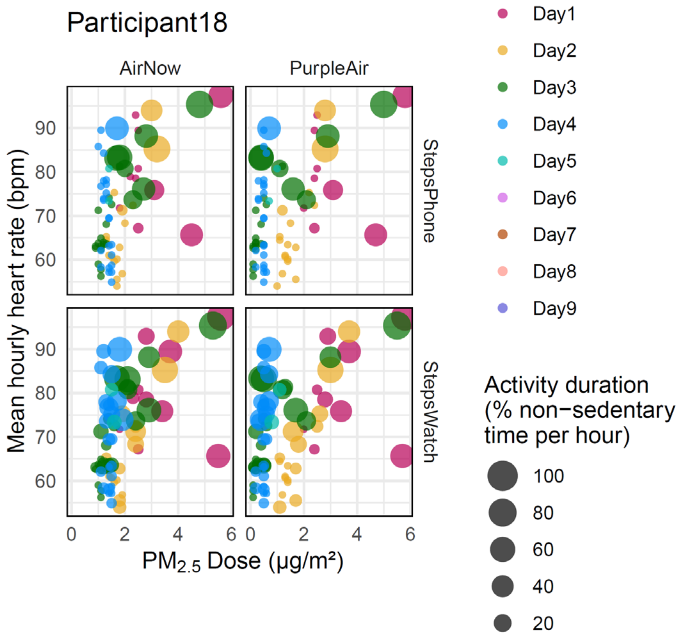

Hourly inhaled doses of $PM_{2.5}$ plotted against mean hourly hear rate where size of each data point indicates activity duration calculated as % non-sedentary time per hour. Note the high degree of inter-day variability observed in many participants.

***Supplementary Figure 11 – Diurnal variability of heart rate***

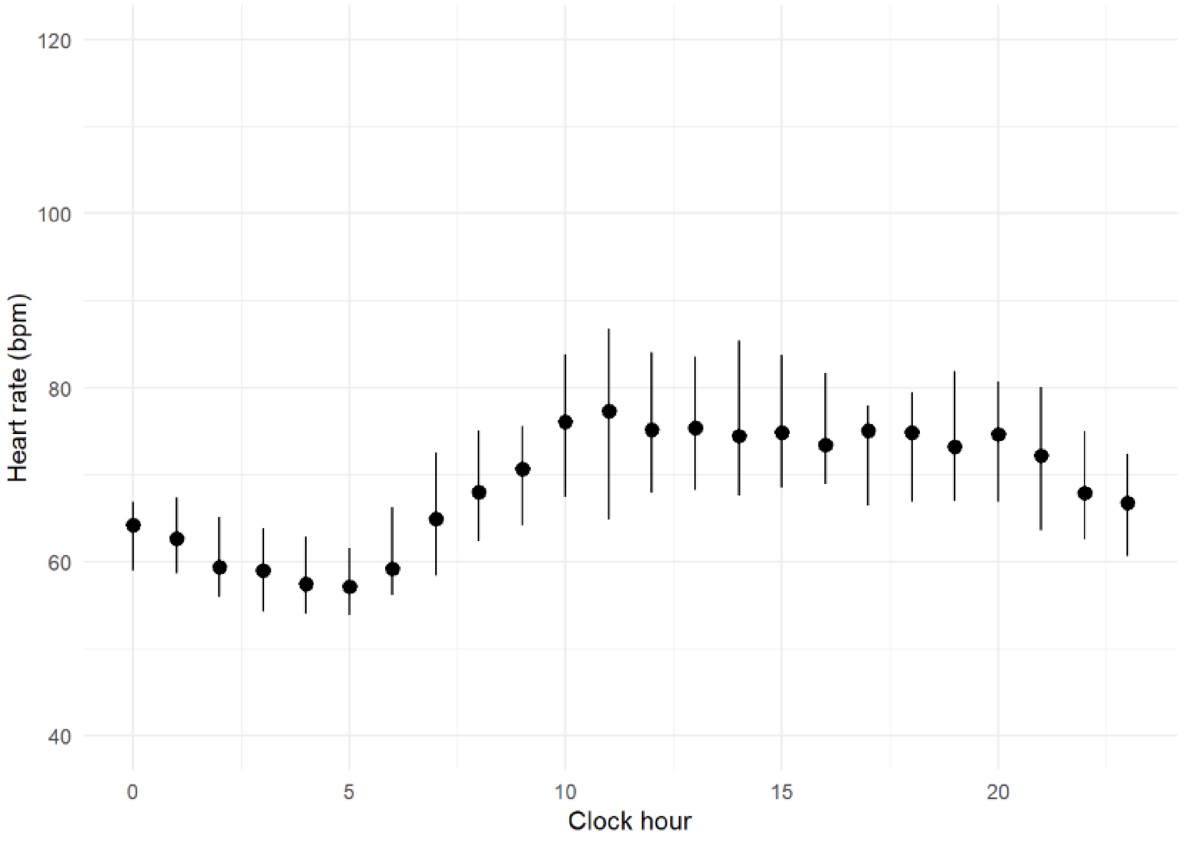

Distributions of heart rates, measured by Apple watch, by hour of day, combined across all study participants. Note: median values are shown as dots, 25th and 75th percentiles shown as lines

#### *Supplementary Figure 12 – Time-specific variability in exposure metrics*

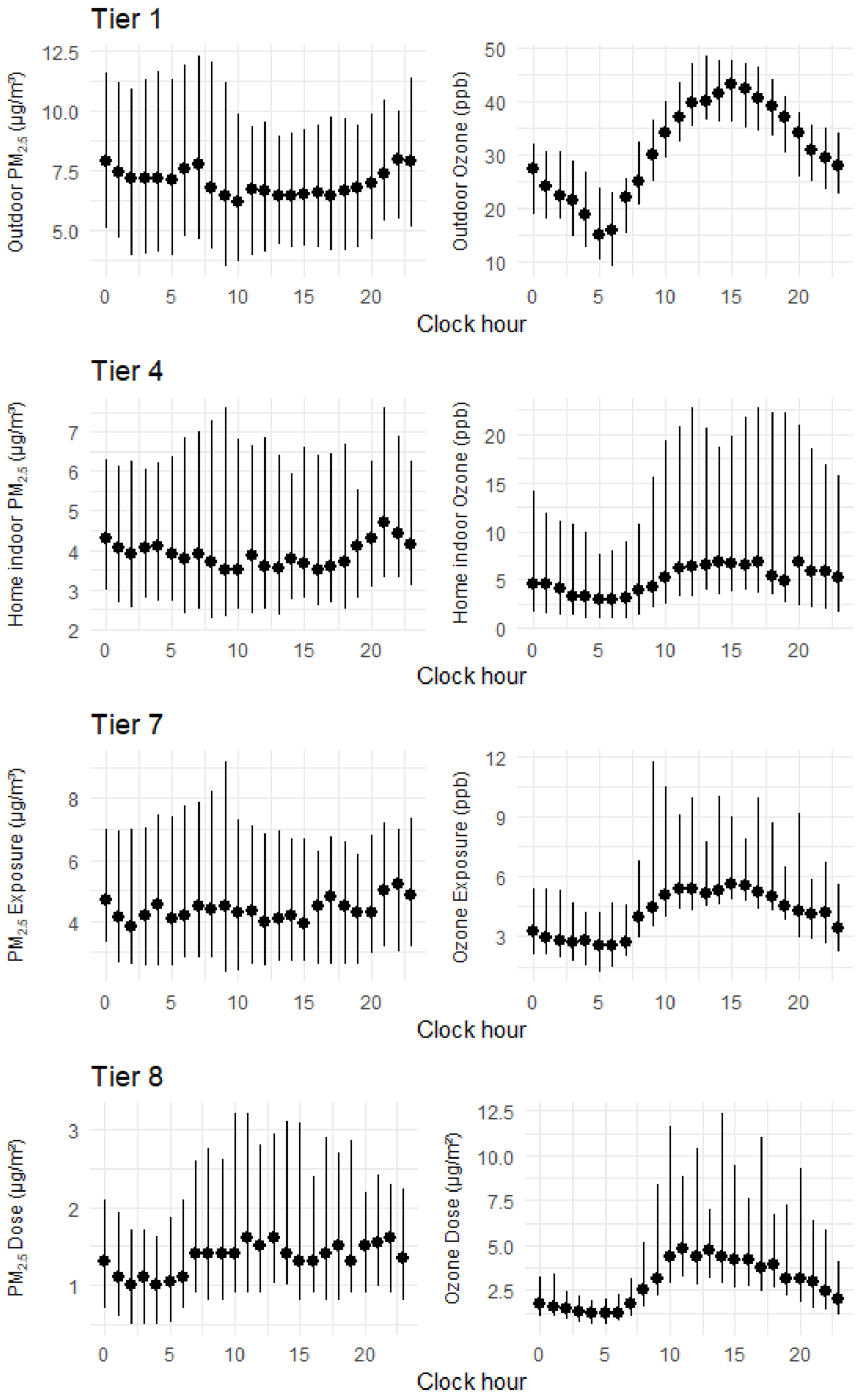

Hourly distributions of $PM_{2.5}$ and ozone metrics from various tiers in the TracMyAir framework (Tier 1 – outdoor concentrations, Tier 4 – indoor concentrations, Tier 7 – exposure concentrations, Tier 8 – heart rate derived inhaled doses), combined across all study participants. Note: median values are shown as dots, 25th and 75th percentiles shown as lines.

*Supplementary Figure 13 – Time-specific variability in inhaled dose metrics*

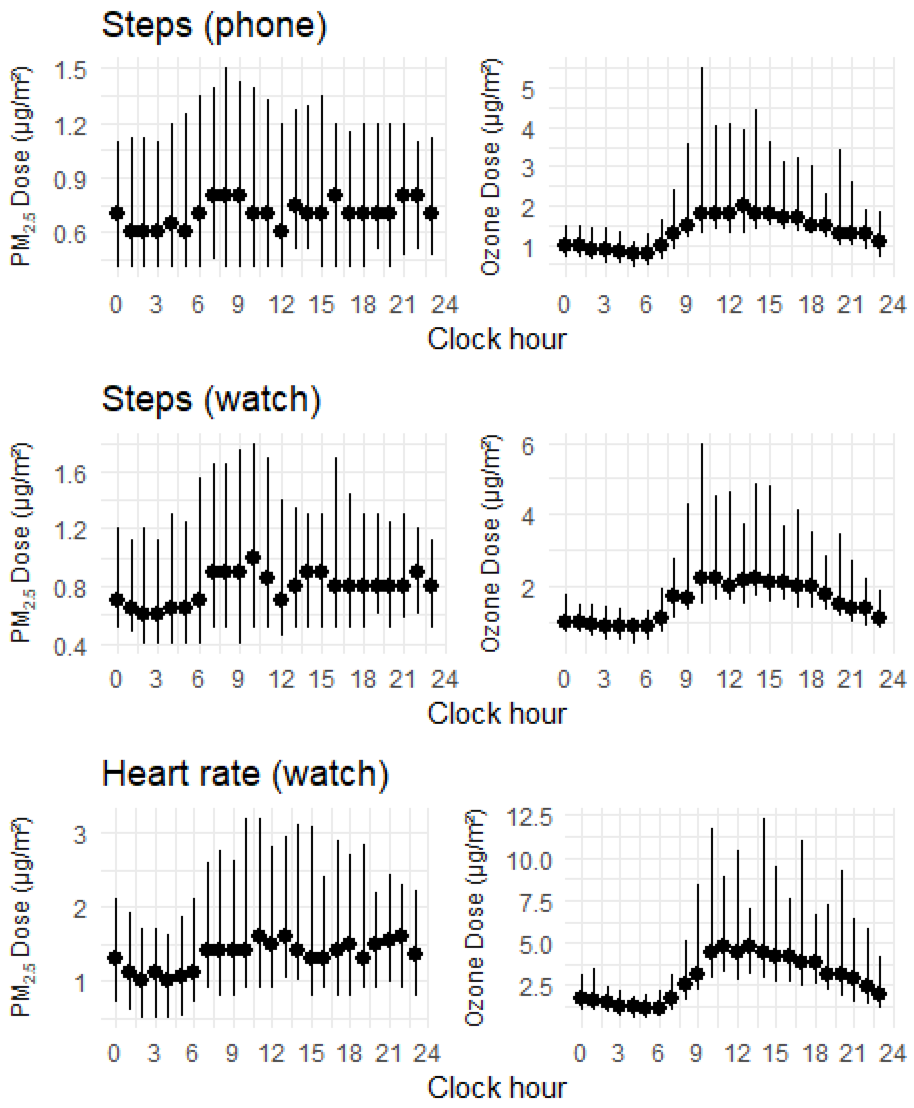

Distributions of inhaled dose calculated using activity from iPhone step counts, Apple watch step counts, and Apple watch heart rates by hour of day for $PM_{2.5}$ and ozone, combined across all study participants. Note: median values are shown as dots, 25th and 75th percentiles shown as lines.

***Supplementary Figure 14 – Air quality in the socioeconomic context***

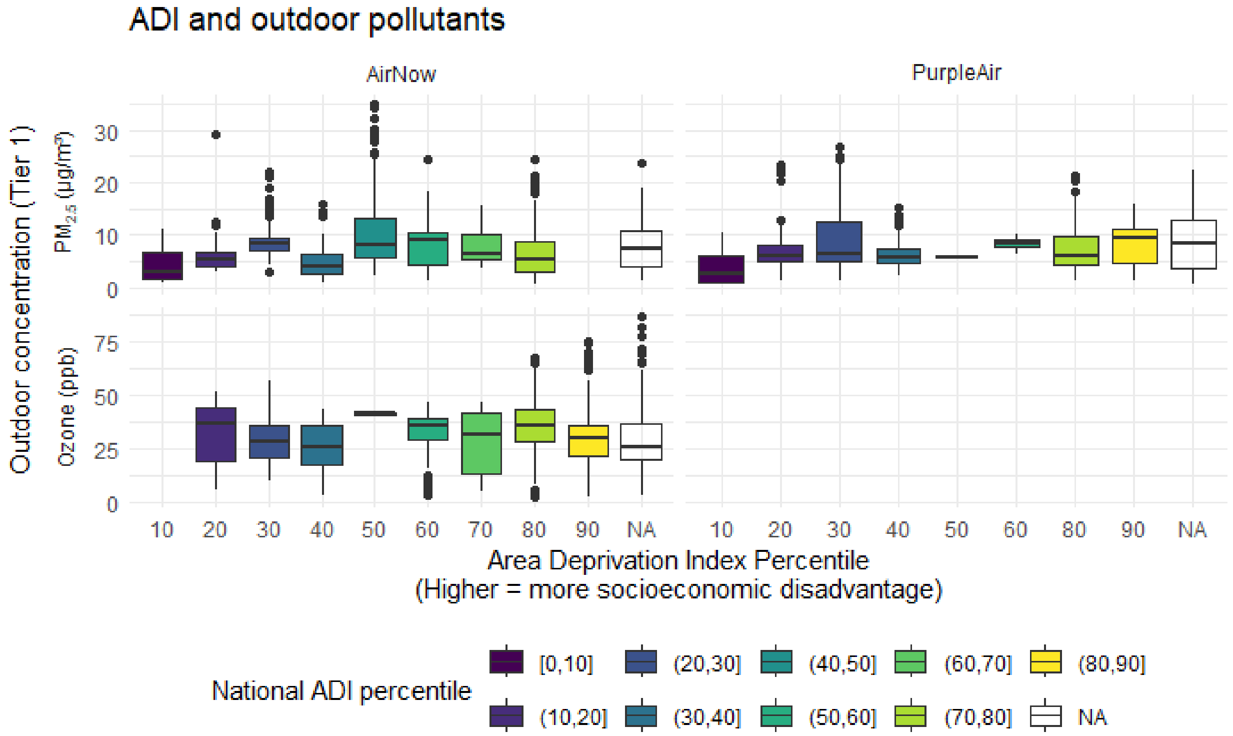

Comparison of outdoor $PM_{2.5}$ and Ozone concentrations to national Area Deprivation Index (ADI) percentiles. Higher ADI percentiles indicate higher degree of socioeconomic disadvantage. NA values are from census block group lacking an ADI due to one of the following flagged conditions: i) GQ (Group Quarters) - Greater than 33.3% of Housing Units are Group Quarters; ii) PH (Population/Housing) - Population less than 100 and/or fewer than 30 housing units; iii) GQ-PH (Group Quarters and Population Housing) - Both GQ and PH conditions are met; and iv) QDI (Questionable Data Integrity) - Block Groups missing a key demographic factor for ADI construction (suppressed or missing in the ACS data). Boxplot lines identify first quartile, median, and third quartile of each distribution.

***Supplementary Figure 15 - Community-driven outdoor monitoring of PM2.5 levels***

a)

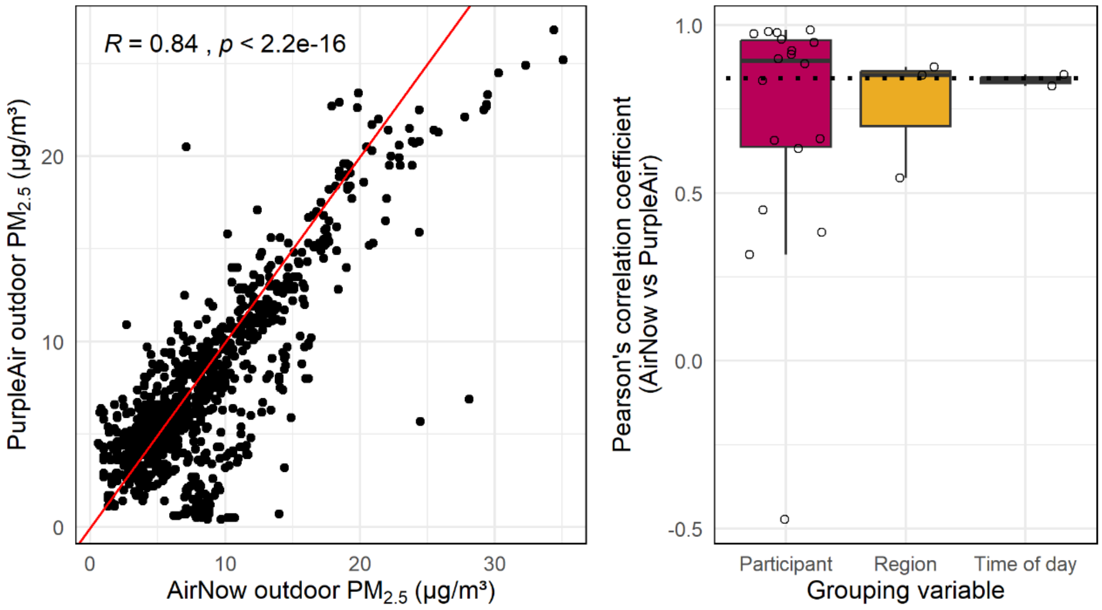

b)

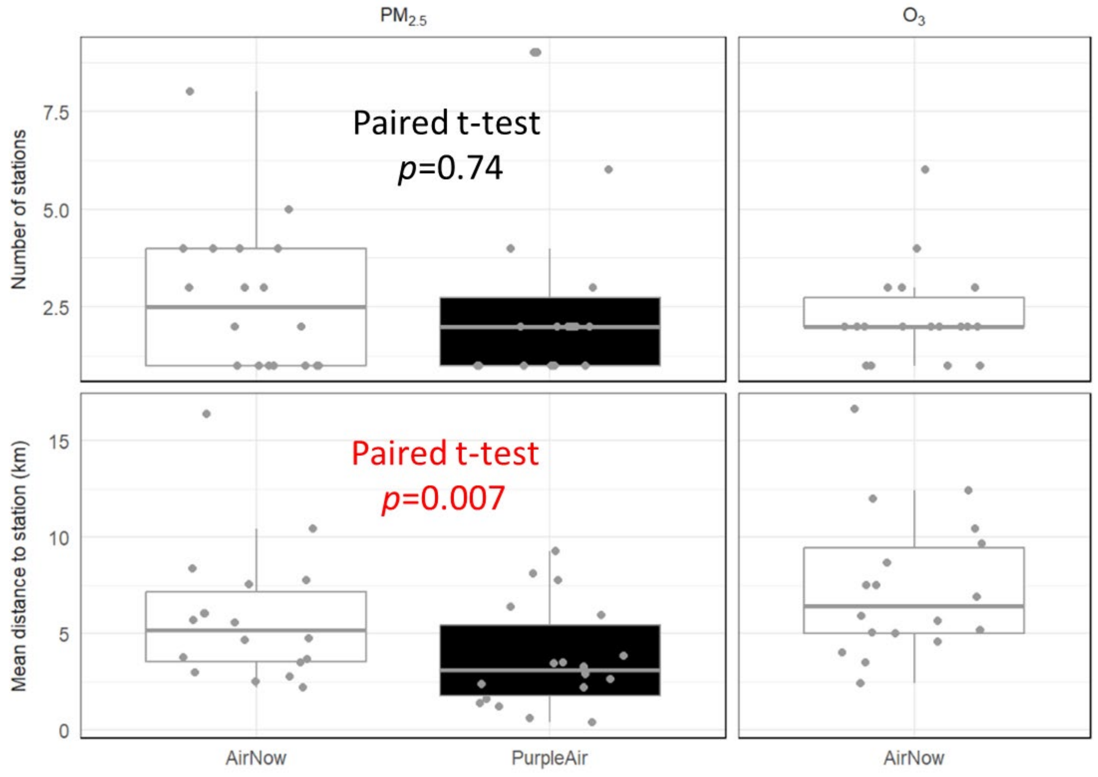

a) Left panel – Scatterplot comparing raw outdoor $PM_{2.5}$ concentrations (Tier 1) as measured by the AirNow (x-axis) and PurpleAir (y-axis) networks, including a simple regression line (red). R value is Pearson's correlation coefficient and p-value is from the

Student's t-test for association. Each point corresponds to a single hour of data from a participant where $PM_{2.5}$ concentrations were measured by both networks. Right panel – distributions of Pearsons's correlation coefficients between AirNow and PurpleAir concentrations when data are grouped either by participant ID, region (Pennsylvania, New Jersey, All others), or time of day (Day, Night). Black dotted line indicates the value of the correlation coefficient from the left panel, where all data are analyzed together. Note that distributions of the grouped coefficients are centered at the value of the ungrouped correlation coefficient. b) Distributions of number of monitoring stations each participant encountered (top) and their average distances from the monitoring stations (bottom) over the course of the study period. Distributions are separated according to monitoring station network (AirNow – white; PurpleAir – black) and the type of molecule measured ($PM_{2.5}$ – left; Ozone – right). Each point corresponds to a single participant. For $PM_{2.5}$ monitoring stations, p-values comparing station metrics between the AirNow and PurpleAir networks are from paired Student's t-tests (paired within study participant).

### *Supplementary Figure 16 - Distributions of exposure metrics from AirNow*

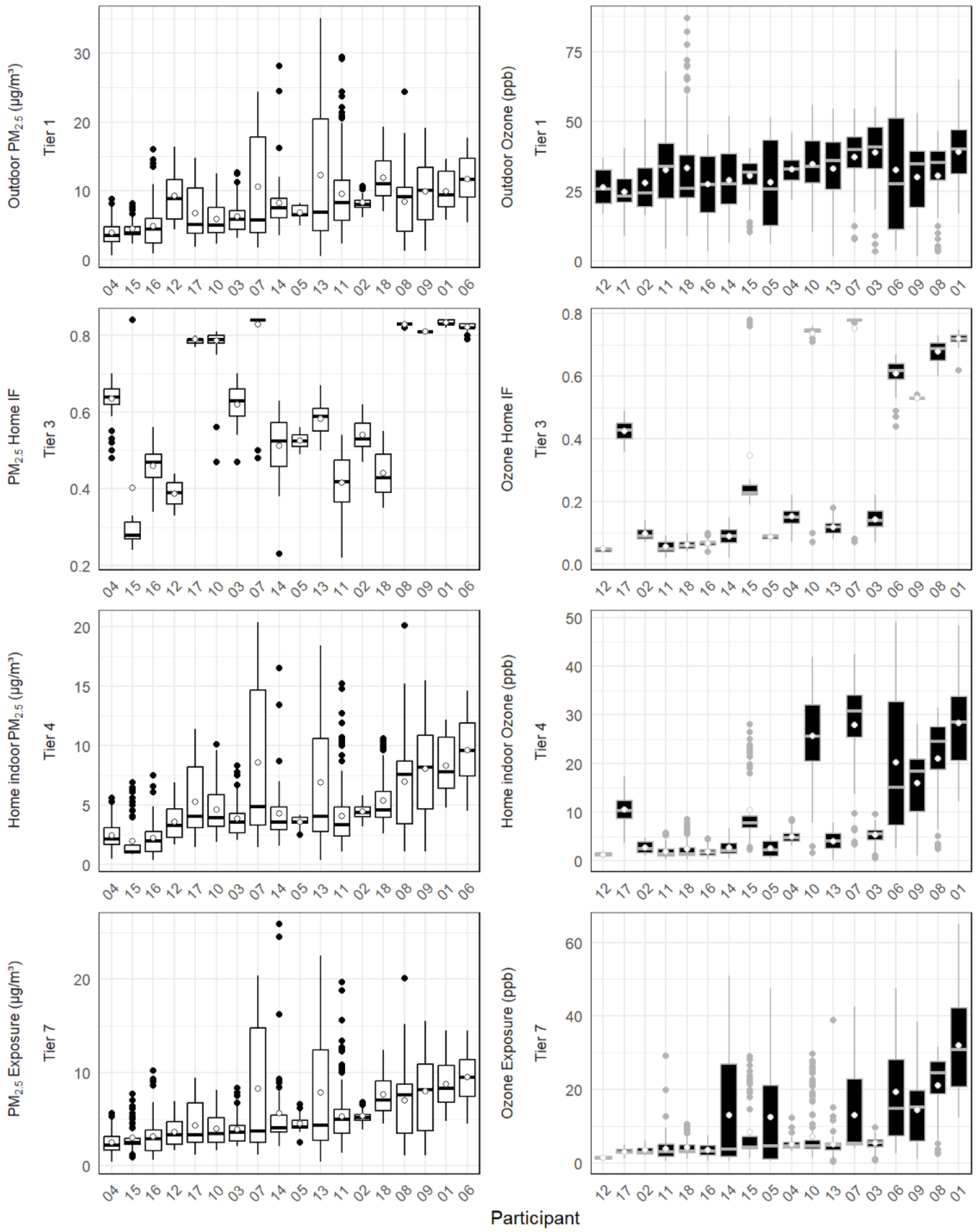

Distributions of exposure metrics (Tier 1: outdoor concentrations, Tier 3: infiltration factors, Tier 4: indoor concentrations, Tier 7: exposure) from TracMyAir sorted by median values for $PM_{2.5}$ and ozone from AirNow monitoring stations for 18 study participants. Boxplot lines identify first quartile, median, and third quartile of each distribution. White dots within boxplots identify distribution means.

***Supplementary Figure 17 - Distributions of exposure metrics from PurpleAir***

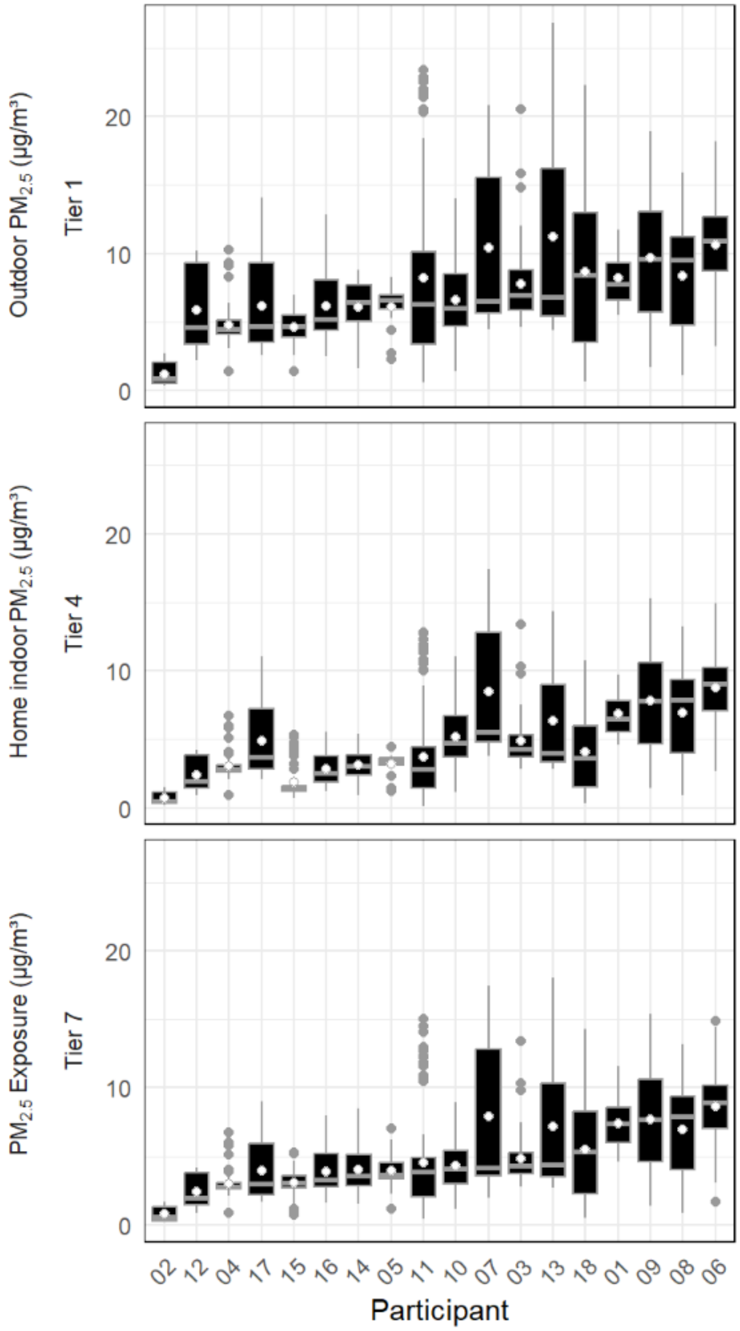

Distributions of Tier 1, 4, and 7 exposure metrics (outdoor concentrations, home indoor, and exposure) for $PM_{2.5}$ from PurpleAir sensors for 18 study participants. Boxplot lines identify first quartile, median, and third quartile of each distribution. White dots within boxplots identify distribution means.

***Supplementary Figure 18 – Study-deployed vs community-embedded PurpleAir sensors***

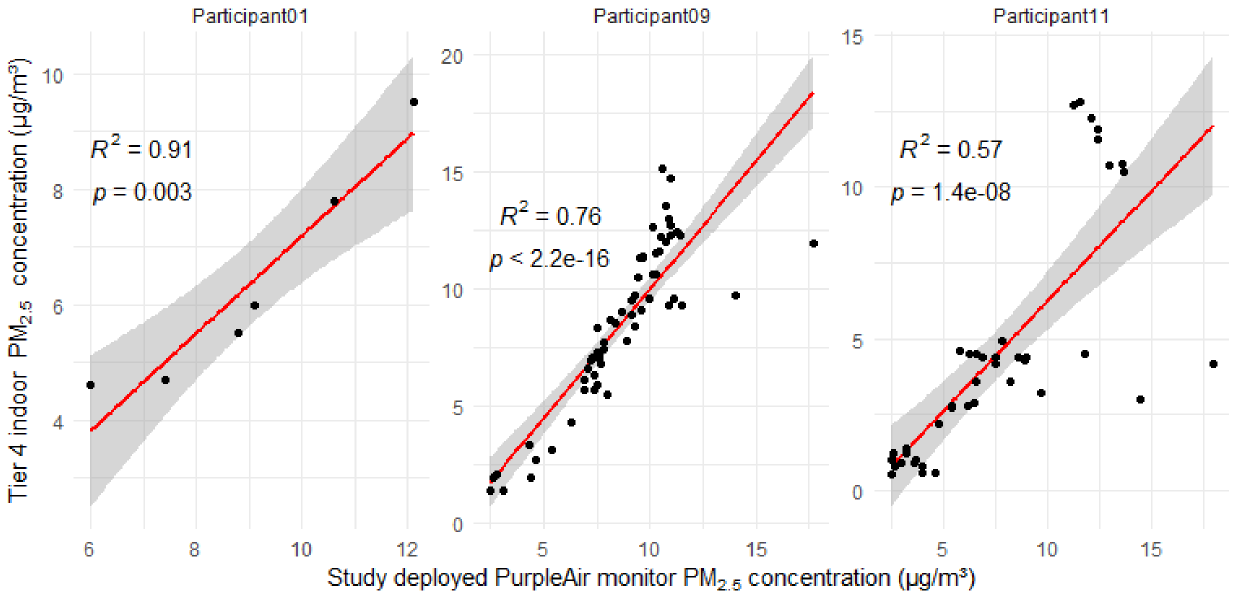

Pearson correlations of indoor $PM_{2.5}$ concentrations measured directly from study-deployed PurpleAir sensors successfully installed for data monitoring in n=3 participants' homes (abscissa) and estimated indoor concentrations (Tier 4) based on data collection from community-embedded PurpleAir sensors (ordinate). Grey areas are the 95% confidence intervals for the regression lines (red). $R^2$ values are Pearson's coefficients of determination and p-values are from the Student's t-test for association (implemented by the cor.test function in base R).

#### *Supplementary Figure 19 - Cohort distributions of modeled exposures to PM2.5 and ozone*

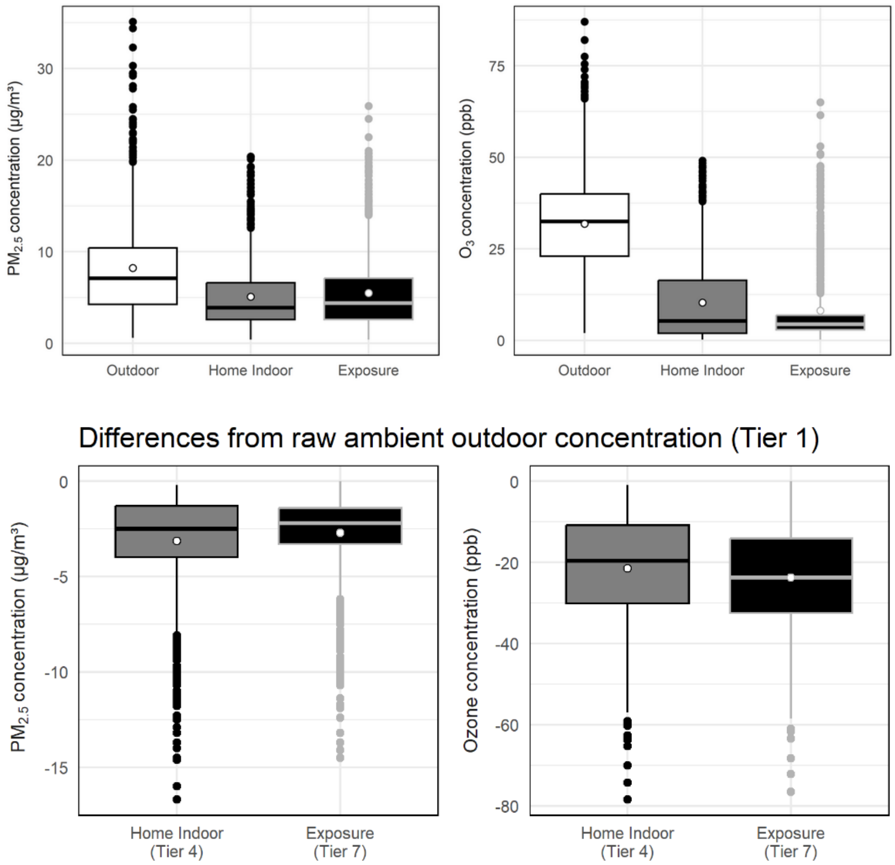

Distributions depicting the difference between home indoor (Tier 4) / exposures (Tier 7) concentrations and outdoor concentrations (Tier 1) for $PM_{2.5}$ (left panels) and ozone (right panels) using AirNow-based estimates as model inputs. Bottom two panels represent the distributions of the Tier 4 and Tier 7 concentrations as differences from the Tier 1 outdoor concentrations. Boxplot lines identify first quartile, median, and third quartile of each distribution. White dots within boxplots identify distribution means.

### ***Supplemental Tables***

***Table S 1 - TracMyAir output variables – infiltration concentrations and exposure***

| | |
|---|---|
| | Home air exchange rate (/hr) |
| | Home PM2.5 infiltration factor |
| | Home indoor PM2.5 level (μg/m³) |
| | Personal PM2.5 exposure factor |
| | Home ozone infiltration factor |
| | Home indoor ozone level (ppb) |
| | Personal ozone exposure factor |
| | Other building PM2.5 infiltration factor |
| | PurpleAir outdoor temperature (ºC) |
| | PurpleAir indoor PM2.5 level (μg/m³) |
| | Non-ambient indoor PM2.5 level (μg/m³) |
| | Non-ambient indoor PM2.5 exposure (μg/m³) |
| | PurpleAir indoor temperature (ºC) |
| | Regression indoor PM2.5 level (μg/m³) |
| | Number of samples used for regression |
| | Home air exchange rate (/hr) |

***Table S 2 - TracMyAir output variables – Exposure Metrics for Epidemiologic Analysis***

Hourly PM2.5 outdoor concentrations (μg/m³)
Hourly ozone concentrations (ppb)
Hourly PM2.5 Home-In concentrations (μg/m³) – ambient, non-ambient, total
Hourly ozone Home-In concentrations (ppb) – ambient
Hourly PM2.5 exposure (μg/m³) in each ME
Hourly ozone exposure (ppb) in each ME
Hourly time spent in each ME (% of hour)
Hourly PM2.5 dose (μg/m²) in each ME & activity level from step counts (2), heart rate
Hourly ozone dose (μg/m²) in each ME & activity level from step counts (2) , heart rate
Hourly time spent at each physical activity level (% of hour)

***Table S 3 - TracMyAir output variables – Concentrations and exposure***

Subject ID
Start time - UTC and Local
PM2.5 monitor location (lat/lon) and distance to monitor (km)
Outdoor PM2.5 concentration (μg/m³)
Ozone monitor location (lat/lon) and distance to monitor (km)
Outdoor ozone concentration (ppb)
Weather station ID, location (lat/lon) and distance to monitor (km)
Outdoor temperature (ºC)
Outdoor wind speed (km/hr)
PM2.5 exposure (μg/m³)
Ozone exposure (ppb)
ME= inside vehicles: Time spent (%), PM2.5 exposure (μg/m³), Ozone exposure (ppb)
ME= indoors at home: Time spent (%), PM2.5 exposure (μg/m³), Ozone exposure (ppb)
ME= outdoors at home: Time spent (%), PM2.5 exposure (μg/m³), Ozone exposure (ppb)
ME= indoors at work: Time spent (%), PM2.5 exposure (μg/m³),Ozone exposure (ppb)
ME= outdoors at work: Time spent (%), PM2.5 exposure (μg/m³),Ozone exposure (ppb)
ME= indoors at unknown: Time spent (%), PM2.5 exposure (μg/m³),Ozone exposure (ppb)

***Table S 4 - TracMyAir output variables – Dose***

PM2.5 dose (μg/m²)
PM2.5 dose [inside vehicles] (μg/m²)
PM2.5 dose [indoors at home] (μg/m²)
PM2.5 dose [outdoors at home] (μg/m²)
PM2.5 dose [indoors at work] (μg/m²)
PM2.5 dose [outdoors at work] (μg/m²)
PM2.5 dose [indoors at unknown] (μg/m²)
Ozone dose (μg/m²)
Ozone dose [inside vehicles] (μg/m²)
Ozone dose [indoors at home] (μg/m²)
Ozone dose [outdoors at home] (μg/m²)
Ozone dose [indoors at work] (μg/m²)
Ozone dose [outdoors at work] (μg/m²)
Ozone dose [indoors at unknown] (μg/m²)

Sedentary intensity time (%)
PM2.5 dose [Sedentary] (μg/m²)
Ozone dose [Sedentary] (μg/m²)
Light intensity time (%)
PM2.5 dose [Light] (μg/m²)
Ozone dose [Light] (μg/m²)
Moderate intensity time (%)
PM2.5 dose [Moderate] (μg/m²)
Ozone dose [Moderate] (μg/m²)
Vigorous intensity time (%)
PM2.5 dose [Vigorous] (μg/m²)
Ozone dose [Vigorous] (μg/m²)

***Table S 5 - TracMyAir individual-level exposure metrics for ambient PM2.5 and ozone***

Tiers of individual-level exposure metrics for ambient $PM_{2.5}$ and ozone in TracMyAir: personal outdoor concentrations ($C_{out}$, Tier 1), residential air exchange rates (AER, Tier 2), infiltration factors ($F_{inf\_home}$, Tier 3), indoor concentrations ($C_{in\_home}$, Tier 4), personal exposure factors ($F_{pex}$, Tier 5), time spent in ME ($T_{ME}$, Tier 6), exposures (E, Tier 7), and inhaled doses (D, Tier 8).

| Tier # | Description, equation |
|---|---|
| Tier 1 | ($C_{out}$) two sets of exposure metrics: 1) AirNow $PM_{2.5}$ and ozone and 2) PA $PM_{2.5}$ |
| Tier 2 | (AER) determined from home building characteristics and weather using LBLX model |
| Tier 3 | ($F_{inf\_home}$) $F_{inf\_home} = P\ AER/(AER + k_r + k_c)$ |
| Tier 4 | ($C_{in\_home}$) $C_{in} = F_{inf}\ C_{out}$ |
| Tier 5 | ($F_{pex}$) $F_{pex} = f_{in\_home}\ F_{inf\_home} + (f_{in\_w} + f_{in\_sch} + f_{in\_cl} + f_{in\_other}) F_{inf\_other\_bldg} + f_{in\_vehicle}\ F_{inf\_vehicle}$ |
| Tier 6 | ($T_{ME}$) from phone geolocations, speed, and building boundaries using MicroTrac |
| Tier 7 | (E) $E = F_{pex}\ C_{out}$ ($E_1 = f_{in\_home}\ F_{inf\_home}\ C_{out}$, $E_2 = f_{in\_work}\ F_{inf\_other\_bldg}\ C_{out}$) |
| Tier 8 | (D) $DS_{ij} = E_i\ MVS_j\ AT / BSA$ |

***Table S 6 – Area deprivation index (ADI) by station and measurement***

| Data Source | Molecule | Stations with valid ADI | Stations with NA ADI | Measurements from valid ADI | Measurements from NA ADI | NA by PH | NA by GQ PH | NA by QDI |
|---|---|---|---|---|---|---|---|---|
| **AirNow** | PM2.5 | 14 | 3 | 797 | 466 | 457 | 9 | 0 |
| **AirNow** | Ozone | 12 | 2 | 1153 | 110 | 110 | 0 | 0 |
| **OpenAQ** | PM2.5 | 10 | 3 | 361 | 103 | 100 | 0 | 3 |
| **OpenAQ** | Ozone | 10 | 4 | 337 | 127 | 122 | 2 | 3 |
| **PurpleAir** | PM2.5 | 23 | 1 | 1078 | 91 | 91 | 0 | 0 |

Breakdown of measurements from stations with valid ADI values and those with NA for ADI rank. Separate counts are included for each ADI flag. The key for the different ADI flag meanings are as follows: i) GQ (Group Quarters) - Greater than 33.3% of Housing Units are Group Quarters; ii) PH (Population/Housing) - Population less than 100 and/or fewer than 30 housing units; iii) GQ-PH (Group Quarters and Population Housing) - Both GQ and PH conditions are met; and iv) QDI (Questionable Data Integrity) - Block Groups missing a key demographic factor for ADI construction (suppressed or missing in the ACS data).

### *Table S 7 - Percent of measurements spent in each microenvironment (ME)*

Percent of measurements spent in each micro environment (ME)

Averaged across all participants

| | AirNow | | | PurpleAir | | |
|---|---|---|---|---|---|---|
| | All | Day | Night | All | Day | Night |
| **Indoors vs Outdoors** | | | | | | |
| Indoors | 91.69% | 88.13% | 99.88% | 90.33% | 87.00% | 99.88% |
| Outdoors | 5.79% | 8.30% | 0.00% | 7.14% | 9.44% | 0.00% |
| Vehicle | 0.59% | 0.83% | 0.00% | 0.58% | 0.83% | 0.00% |
| Mixed | 1.94% | 2.75% | 0.12% | 1.95% | 2.73% | 0.12% |
| **Home vs Work** | | | | | | |
| Home | 52.23% | 48.28% | 60.93% | 51.47% | 47.95% | 60.66% |
| Work | 3.28% | 4.66% | 0.00% | 4.19% | 5.34% | 0.00% |
| Other | 41.83% | 43.28% | 38.95% | 41.63% | 42.89% | 39.22% |
| Vehicle | 0.59% | 0.83% | 0.00% | 0.58% | 0.83% | 0.00% |
| Mixed | 2.07% | 2.95% | 0.12% | 2.13% | 2.99% | 0.12% |
| **Specific MEs** | | | | | | |
| Indoors Home | 49.26% | 43.98% | 60.93% | 47.97% | 43.11% | 60.66% |
| Outdoors Home | 2.24% | 3.25% | 0.00% | 2.22% | 3.24% | 0.00% |
| Indoors Work | 0.20% | 0.28% | 0.00% | 0.25% | 0.34% | 0.00% |
| Outdoors Work | 3.08% | 4.37% | 0.00% | 3.93% | 5.00% | 0.00% |
| Indoors Unknown | 14.58% | 14.88% | 13.82% | 14.16% | 14.40% | 13.60% |
| Indoors Other | 27.25% | 28.40% | 25.12% | 27.47% | 28.50% | 25.62% |
| Inside Vehicles | 0.59% | 0.83% | 0.00% | 0.58% | 0.83% | 0.00% |
| Mixed | 2.81% | 4.00% | 0.12% | 3.40% | 4.59% | 0.12% |

Values in table columns do not sum to 100% because they are means calculated across all participants.

### ***Supplemental References***

1 Hossain, M. S., Che, W., Frey, H. C. & Lau, A. K. H. Factors affecting variability in infiltration of ambient particle and gaseous pollutants into home at urban environment. *Building and Environment* **206**, 108351 (2021). https://doi.org:https://doi.org/10.1016/j.buildenv.2021.108351

2 Breen, M. S., Xu, Y., Christopher Frey, H., Breen, M. & Isakov, V. Microenvironment Tracker (MicroTrac) model to estimate time-location of individuals for air pollution exposure assessments: model evaluation using smartphone data. *J Expo Sci Environ Epidemiol* **33**, 407-415 (2023). https://doi.org:10.1038/s41370-022-00514-w

3 Klepeis, N. E. *et al.* The National Human Activity Pattern Survey (NHAPS): a resource for assessing exposure to environmental pollutants. *J Expo Anal Environ Epidemiol* **11**, 231-252 (2001). https://doi.org:10.1038/sj.jea.7500165
